# Clinical impact and cost-effectiveness of updated 2023/24 COVID-19 mRNA vaccination in high-risk populations in the United States

**DOI:** 10.1101/2024.11.15.24317369

**Authors:** Keya Joshi, Mariia Dronova, Ewelina Paterak, Van Hung Nguyen, Hagit Kopel, James Mansi, Nicolas Van de Velde, Ekkehard Beck

## Abstract

**Introduction:** In the post-pandemic era, people with underlying medical conditions continue to be at increased risk for severe COVID-19 disease, yet COVID-19 vaccination uptake remains low. This study estimated the clinical and economic impact of updated 2023/24 Moderna COVID-19 vaccination among high-risk adults versus no updated vaccination and versus updated Pfizer/BioNTech vaccination.

**Methods:** A static Markov model was adapted for high-risk adults, including immunocompromised (IC), chronic lung disease (CLD), chronic kidney disease (CKD), cardiovascular disease (CVD), and diabetes mellitus (DM) populations in the United States.

**Results:** Vaccination with the updated Moderna vaccine at current coverage rates was estimated to prevent considerable COVID-19 hospitalizations in CLD (101,309), DM (97,358), CVD (47,830), IC (14,834) and CKD (13,558) populations versus no updated vaccination. Vaccination also provided net medical cost savings of $399M–2,129M (healthcare payer) and $457M–2,531M (societal perspective), depending on population. The return-on-investment was positive across all conditions ($1.10–$2.60 gain for every $1 invested). Healthcare savings increased with a relative 10% increase in current vaccination coverage ($439M–$2,342M), and from meeting US 2030 targets of 70% coverage ($1,096M–$5,707M). Based on higher vaccine effectiveness observed in real-world evidence studies, updated Moderna vaccination was estimated to prevent additional COVID-19 hospitalizations in DM (13,105), CLD (10,359), CVD (6,241), IC (1,979), and CKD (942) versus Pfizer/BioNTech’s updated vaccine, with healthcare payer and societal cost savings, making it the dominant strategy. Healthcare savings per patient vaccinated with Moderna versus Pfizer/BioNTech’s updated vaccine were $31-59, depending on population. Results were robust across sensitivity/scenario analyses.

**Conclusions:** Updated 2023/24 Moderna COVID-19 vaccination was estimated to provide significant health benefits through prevention of COVID-19 in high-risk populations, and cost-savings to healthcare payers and society, versus no vaccination and updated Pfizer/BioNTech vaccination. Increasing current low COVID-19 vaccination coverage rates was estimated to be cost-saving while preventing many more severe infections and hospitalizations in these high-risk populations.

**Key Summary Points:** *Why carry out this study?:*

- In the US, people with underlying medical conditions continue to be at high risk of severe COVID-19, yet vaccination rates are low.
- The CDC recommends an updated 2024/25 COVID-19 vaccination for everyone aged >6 months.
- The objective of this study was to estimate the clinical benefits and cost-effectiveness of updated 2023/24 Moderna COVID-19 vaccination in people with high-risk conditions, versus no updated vaccination, and versus updated Pfizer/BioNTech COVID-19 vaccination.

*What was learned from the study?:*

- COVID-19 vaccination with the updated Moderna mRNA vaccine of people with underlying medical conditions at high-risk of severe COVID-19 was cost-saving versus no updated vaccination. It also provided more health gains with cost savings versus Pfizer/BioNTech, making it the dominant strategy.
- For every $1 spent on vaccination, the updated Moderna vaccination provided a return-on-investment of $1.10–$2.60 versus no updated vaccination, depending on the high-risk population. Healthcare cost savings were $31-59 per patient vaccinated with Moderna’s versus Pfizer/BioNTech’s updated vaccination, depending on the high-risk population.
- A relative 10% increase in vaccination coverage rates prevented 10% more hospitalizations and deaths, and increased healthcare and societal cost savings in all high-risk populations, with the potential for significant health and financial benefits with greater vaccine coverage rates.

## 1. Introduction

Even in the post-COVID-19 pandemic era, COVID-19 remains the most frequent cause of respiratory hospitalizations in the United States (US), surpassing both influenza and respiratory syncytial virus in winter 2022/23 and winter 2023/24 [1]. COVID-19 was the 10th leading cause of death in 2023 [2]. The Center for Disease Control and Prevention (CDC) continues to recommend COVID-19 vaccination universally for everyone six months and older, to protect against potentially serious outcomes of COVID-19 [3]. To match circulating variants more closely, the CDC recommended vaccines targeting the XBB1.5 sublineage for vaccination in Fall 2023/24 [4], and updated vaccines targeting the KP.2 sublineage of JN.1 from June 2024 [5].

While all individuals are susceptible to severe outcomes following SARS-CoV-2 infection, the CDC recognizes that certain subgroups of the US population continue to be at higher risk of severe COVID-19 disease. In addition to adults aged ≥65 years, individuals with specific underlying medical conditions such as chronic lung disease (CLD), chronic kidney disease (CKD), cardiovascular disease (CVD), diabetes mellitus (DM), and people who are immunocompromised (IC) were identified as being at higher risk for severe outcomes [6]. For these groups, vaccination remains a priority to prevent morbidity and mortality due to COVID-19 [6]. There are an estimated 176 million (M) US adults (75.4% of the population) with at least one medical condition, 40.3% with ≥2, and 18.5% with ≥3 medical conditions. There are an estimated 129M adults <65 years of age with at least one risk factor that places them at increased risk of COVID-19 [7]. Moreover, studies have found that the risk of severe COVID-19 outcomes is higher in people with multiple underlying conditions [8].

COVID-19 hospitalization costs [9] and medical costs in the year following acute infection [10] were shown to be higher in patients with underlying medical conditions. Thus, COVID-19 can contribute to increasing the already high healthcare costs related to underlying conditions e.g., around $70 billion for CLD (asthma and chronic obstructive pulmonary disease) [11]; $87.2 billion for Medicare beneficiaries with CKD [12]; $400 billion for CVD risk factor care (projected to triple by 2050) [13]; and $306.6 billion for DM medical costs [14]. The 2023/24 COVID-19 vaccination coverage in the general population was low at 15.6% among 18–49-year-olds, 24.6% among 50–64-year-olds, and 37.4% among ≥65-year-olds [15], thus many high-risk individuals in the US remain vulnerable to severe COVID-19.

Three COVID-19 vaccines are currently available in the US: mRNA vaccines from Moderna and Pfizer/BioNTech (both recommended for individuals aged ≥6 months) and a protein-based vaccine from Novavax (recommended for individuals aged ≥12 years). There are differences between the Moderna and Pfizer/BioNTech mRNA vaccines (e.g., formulation, delivery system, and dosage [16]). Real-world evidence studies comparing the COVID-19 mRNA vaccines (Moderna mRNA-1273 and Pfizer-BioNTech BNT162b2) in populations with underlying medical conditions found differences in vaccine effectiveness between the two vaccines e.g., in studies of bivalent original/Omicron BA4-5 containing vaccines [17, 18], and in a recent systematic literature review and GRADE meta-analysis, which showed significantly reduced risks with the mRNA-1273 versus BNT162b2 vaccine, of SARS-CoV-2 infection (RR 0.85 [95% CI, 0.79-0.92], severe SARS-CoV-2 infection (RR 0.83 [95% CI, 0.78–0.89]), COVID-19 related hospitalization (RR 0.88 [95% CI, 0.82–0.94]), and COVID-19 related death (RR 0.84 [95% CI, 0.76–0.93]) [19].

As SARS-CoV-2 continues to evolve, it is important for decision-makers, public health officials, and healthcare practitioners to understand the clinical and economic benefits of current vaccination programs as well as the benefits associated with increasing vaccination rates, to optimize clinical management by minimizing disease burden at the individual and system level, especially for those with high-risk conditions.

The objective of this analysis was to estimate the clinical and economic impact of updated 2023/24 COVID-19 vaccination, by comparing vaccination with the updated Moderna COVID-19 vaccine versus no updated vaccination in adults ≥18 years with underlying medical conditions (i.e., IC, CVD, DM, CKD, or CLD) associated with increased risk for severe COVID-19. In addition, the potential cost-effectiveness of vaccination with Moderna’s versus Pfizer/BioNTech’s updated vaccines was assessed, assuming comparable differences in VE as those observed [17]. A previously published economic model [20, 21] was updated for this analysis.

## 2. Methods

A published static Markov model developed in Excel [20, 21] was adapted for the US, to assess the cost-effectiveness of updated Moderna COVID-19 vaccination versus no vaccination with any 2023/24 updated vaccine in a primary analysis, in high-risk adults previously diagnosed with the following conditions: IC or had CLD, DM, CVD, or CKD (previously defined [22–27]).

Vaccination and healthcare costs were included (healthcare payer perspective), as well as lost productivity costs (societal perspective). The model time horizon was one year, from September 1^st^, 2023, to August 31^st^, 2024. In a secondary analysis, the clinical impact and cost-effectiveness of vaccination with Moderna’s versus Pfizer/BioNTech’s updated vaccines was compared.

### 2.1. Model overview

The cohort in the static Markov model began in the ‘Well’ health state and had a given risk of symptomatic infection each month based on the projected monthly COVID-19 incidence. The risk of infection was lower for those vaccinated. Progression pathways through the model are illustrated in the decision tree (**Fig. 1**). COVID-19 deaths were assumed to only occur in hospitalized cases, and all infected individuals could develop long COVID.

**Fig 1.**
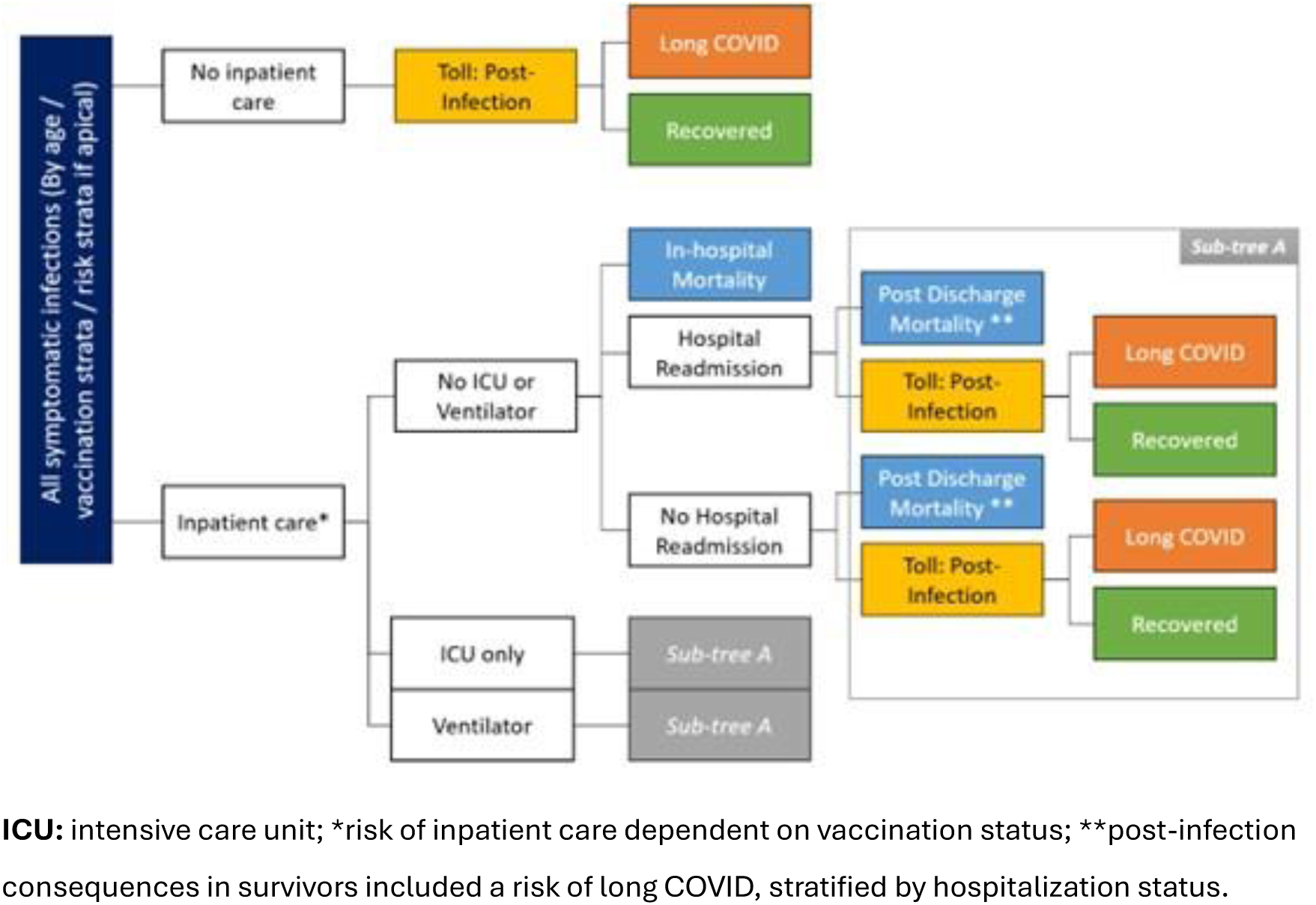
Decision tree model overview based on [21]

### 2.2. Input parameters and model assumptions

#### 2.2.1. Population

The model population included US adults (≥18 years) with highly prevalent (including among young adults) underlying conditions associated with higher risk for severe COVID-19 outcomes i.e., CLD, DM, CVD, CKD and IC populations [17] (Table S1). Each population was assessed separately, as adults may have multiple high-risk conditions [7]. The population sizes were as follows for individuals with: CLD 37.0M [22]; DM 29.3M [23]; CVD 16.6M [24]; IC 7.0M [25]; and CKD 3.7M based on adults diagnosed/aware of CKD, assuming that 90% of an estimated 37M US adults living with CKD are undiagnosed, or not aware of having CKD [26, 27]. The distribution across model age groups was based on single-year US general population estimates [28] and prevalence data [25, 29].

#### 2.2.2. Incidence

Monthly symptomatic COVID-19 incidence (**Table S2**) was informed by a dynamic Susceptible-Exposed-Infected-Recovered (SEIR) model developed for the US, calibrated to ensure the model predicted a valid number of infections [30].

#### 2.2.3. Vaccine effectiveness and coverage

This analysis for updated vaccination in 2023/24 considered the XBB1.5 vaccine, however, it should be noted that the 2023/24 COVID-19 vaccines and prior formulations are no longer authorized for use in the US.

Initial vaccine effectiveness (VE) of the updated Moderna COVID-19 XBB1.5 vaccine in high-risk populations was based on analysis of claims data from September 12^th^ to December 31^st^, 2023 [31]. After the first month of vaccination, monthly linear waning rates for protection against infection and hospitalization were applied, assuming the same waning rates of a monovalent booster dose against omicron during an omicron-dominant period, assessed in a meta-analysis [32]. This was in line with previous economic analyses of COVID-19 vaccination in the US [30] and in IC populations in Canada and France [20, 21]. VE against infection was 34.50% (with waning of 4.75% per month) and VE against hospitalization was 58.70% (with waning of 1.37% per month) (**Table S7**).

The VE for the updated Pfizer/BioNTech COVID-19 vaccine was estimated by applying a relative vaccine effectiveness (rVE) to the VE estimates of Moderna’s updated COVID-19 vaccine (**Table S7**). The rVEs were based on a large database analysis comparing the bivalent (original/ Omicron BA.4/BA.5) COVID-19 mRNA vaccines from Moderna (mRNA1273.222) and Pfizer/BioNTech (BNT162b2) [17], which included data in adults with CLD, DM, CVD and IC adults. The same linear VE waning rates were applied for Moderna’s and Pfizer/BioNTech’s vaccines.

The base-case considered one-dose vaccination starting in September 2023. Vaccination coverage (**Table S8**) was based on cumulative observed COVID-19 vaccination coverage between September 24, 2023 and February 24, 2024 (2023/24 season), reported by the CDC [15]. Uptake was assumed to occur over a six-month period between September 2023 and February 2024, based on observed uptake, and was assumed to be the same for both vaccines.

Vaccinated individuals could experience vaccine-related adverse events: grade 3 local (4.90%) or systemic (7.61%) infection-related events, anaphylaxis (0.0005%) and myocarditis/ pericarditis (ages 18–39 only, 0.0018%) [30]. The risk of adverse events was assumed equal for Moderna’s and Pfizer/BioNTech’s updated vaccines, and based on the previous US economic analysis for COVID-19 in the general population [30].

#### 2.2.4. Probabilities

The proportion of patients not hospitalized with outpatient care (41.00%) was based on influenza data (CDC 2019/20 influenza season) [33]. The rate of hospitalization given symptomatic COVID-19 infection in the unvaccinated general population (10.54% for adults) [31] was adjusted for high-risk groups by applying a risk ratio (RR) from data on disease burden in patients with medical conditions [29, 34] i.e., CLD RR of 1.52; DM RR of 2.01; CVD RR of 1.59; IC RR of 1.20; and CKD RR of 1.89. Although the COVID-19 hospitalization rate is expected to increase with age [35], the model conservatively assumed the same hospitalization rate across all age groups (**Supplemental File S1, Table S3**).

For CKD, CLD, CVD and DM patients, the distribution by in-hospital level of care (no ICU or ventilator; ICU only; ventilator) was derived from the CDC COVID-NET hospitalizations tracker in the season 2023/24 [36]. Due to the lack of condition-specific data, the general population estimates were assumed to be applicable for these populations. The distribution for the IC population was estimated by applying a RR to general population CDC data [35, 36] (**Supplemental File S1, Table S3**).

For CVD, DM and IC patients, in-hospital mortality was estimated by applying an RR to the general population mortality due to COVID-19 [36]: RR of 1.23 for DM [37]; RR of 1.62 for CVD [38]; and RR of 1.74 for IC [39]. For CKD and CLD patients, in-hospital mortality rates were informed by CDC data reported for the general population [36], in line with previous studies [40, 41]. It was assumed that in-hospital mortality rates were the same across all levels of care. (**Supplemental File S1, Table S3**). The risk of post discharge mortality was based on previous estimates from a US economic analysis of COVID-19 [30], due to the lack of data in high-risk patients, and assumed to be the same for ICU only and ventilator (**Table S3**).

The risk of hospital re-admission by location of care was estimated by applying a RR to general population data [30, 42] i.e., CLD RR of 1.08 [42]; DM RR of 1.27 [42]; CVD RR of 1.38 [42]; IC RR of 1.15 [43]; and for CKD, date were derived from the United States Renal Data System (USRDS) [44], and assumed to be the same for all levels of care. It was assumed that re-admission rates were the same for all model age groups (**Supplemental File S1, Table S3**).

Post-infection QALY losses and medical costs were assumed to occur in the year following acute infection and were more severe in hospitalized cases and older age groups [30, 45], while productivity losses were assumed for severe long COVID cases. For CLD, CVD, DM and IC patients, the proportion of individuals with long COVID (overall and severe) was based on a previous US economic analysis for COVID-19 [30], due to a lack of data in the high-risk population. For CKD patients, the proportion of people with additional impact from long COVID (e.g., severe long COVID) was adjusted by data from the USRDS [44] (**Table S4**).

The probability of hospitalization due to infection-related myocarditis in high-risk symptomatic patients was assumed to be the same as for the general population, and was based on a previous US economic analysis for COVID-19 [30] (**Table S4**).

#### 2.2.5. Resource use and costs

Direct healthcare costs included vaccination [46, 47] and vaccine administration costs [48], as well as acute outpatient care ($460.57), hospitalization with no ICU or ventilation ($15,089), with ICU only ($27,058) and with ventilator ($71,367), and inpatient follow-up per case ($1,075), adjusted for each high-risk population when data were available. The proportion of patients not hospitalized with outpatient care (41.00%) was estimated using data on influenza from the CDC during the 2019–2020 influenza season [33]. Hospitalization costs for the general population [49] were not adjusted for the CLD population, as no increase in hospitalization costs was reported for patients with COPD [9], and this was assumed to be the same for CLD patients. For CKD, CVD, DM and IC patients, the general population hospitalization costs [49] were adjusted by a relative cost ratio [9] of 1.08 for DM; of 1.11 for CVD; of 1.25 for the IC population; and of 1.64 for CKD. Post-infection costs [10] reflected all medical costs incurred in the post-acute period for high-risk populations and varied by age and outpatient/inpatient status. (**Table S5**).

Indirect lost productivity costs (**Table S5**) were included from a societal perspective, for those actively participating in the labor force, with calculations for the number of lost working days due to vaccine administration, outpatient care, hospitalization, hospitalization recovery and severe long COVID (see **Supplemental File S2** for detailed estimations also considering adjustments for increased productivity losses due to the underlying condition).

#### 2.2.6. QALYs/ utilities

The model applied baseline utility values stratified by age group [50], and utility decrements sourced from 5UM-CDC data [51] and a previous US economic analysis of COVID-19 in the general population [30], assumed to apply to high-risk populations (**Supplemental File S2, Table S6**).

#### 2.2.7. Scenario analyses

Key model inputs were varied in one-way scenario analyses, and the impact on healthcare cost savings and on QALYs saved was assessed. For the comparison versus no updated vaccination: both higher and lower incidence rate scenarios were assessed, using data from the dynamic SEIR model [30] (**Table S2**); and variations in RR for hospitalization (**Table S3**); VE against infection and against hospitalization (**Table S7**); waning of VE against hospitalization (**Table S7**); and inpatient and post-infection costs (**Table S5**) were varied. For the comparison against Pfizer/BioNTech’s updated vaccine, a scenarios assessed the impact of varying the rVE estimates (**Table S7**).

The impact of increasing relative vaccination coverage rates (VCR) by 10% compared with the current coverage (i.e., the observed VCR for the 2023-2024 season), and of achieving the target coverage rate reported for influenza of 70% [52] across all age groups was assessed (**Table S8**).

Following ACIP recommendations, a scenario assessed the impact of a two-dose vaccination versus no updated vaccination, with an additional Spring vaccine dose for high-risk adults aged ≥65 years who received the updated vaccination. Uptake for the Spring dose was assumed between March and April, reaching 10% coverage in April, based on data on the proportion of people who received two doses by April 27, 2024 [15] (**Table S8**).

### 2.3. Compliance with Ethics Guidelines

This analysis is based on previously conducted studies and does not contain any new studies, with human participants or animals, performed by any of the authors.

## 3. Results

### Primary analysis: updated Moderna COVID-19 vaccination versus no updated vaccination

The model population included individuals with CLD (37.0M), DM (29.3M), CVD (16.6M), IC (7.0M), and CKD (3.7M). The number needed to vaccinate to prevent one hospitalization was 117 (CLD), 87 (DM), 112 (CVD), 116 (IC) and 93 (CKD). The model estimated that between September 2023 and February 2024, updated Moderna COVID-19 vaccination prevented 25,668–255,253 symptomatic COVID-19 infections; 13,558–101,309 COVID-19 hospitalizations; 1,870–13,694 COVID-19 deaths, and 6,606–68,663 long COVID cases compared with no updated vaccination (**Fig. 2**).

**Fig 2.**
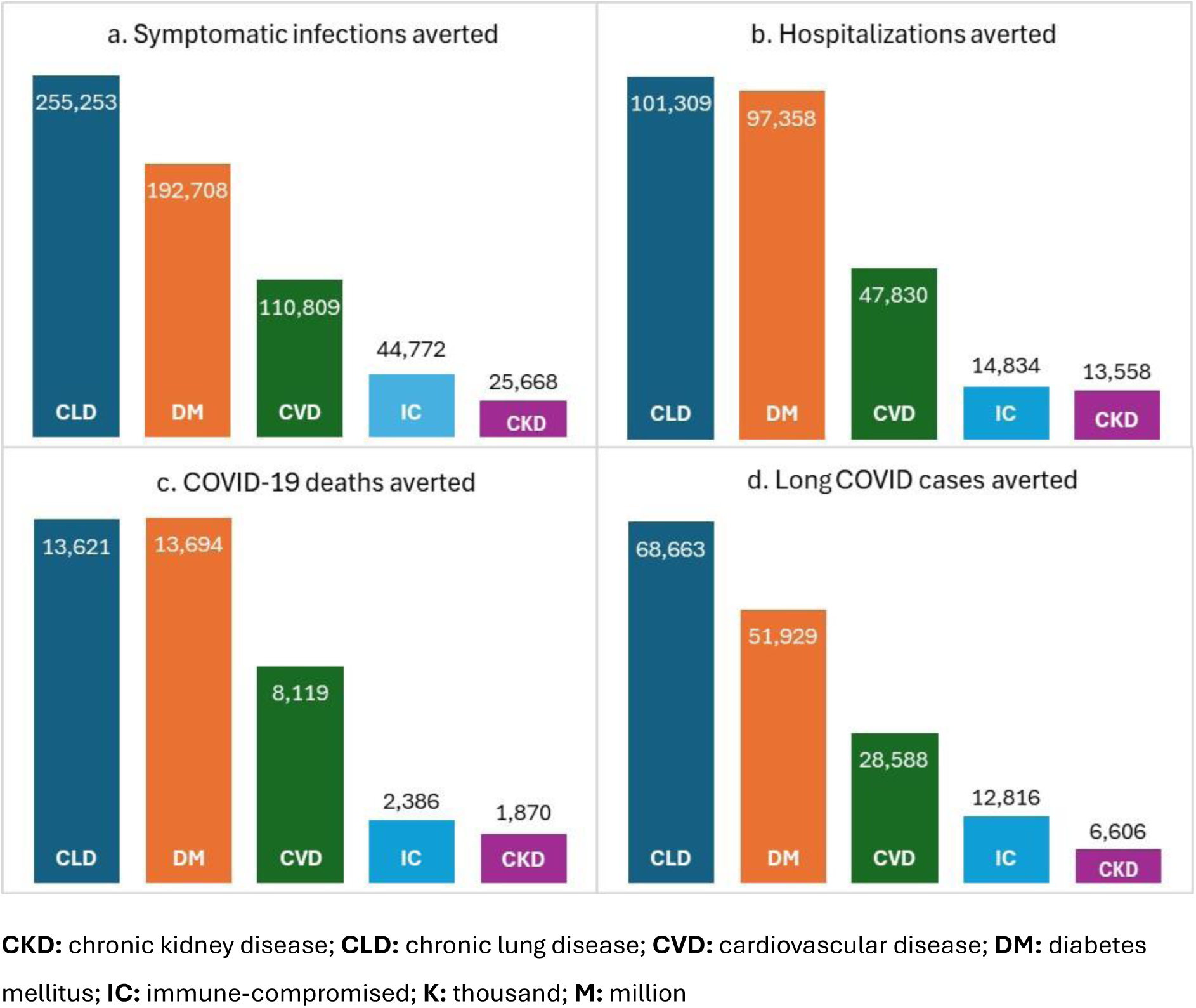
COVID-19 burden averted (numbers, %) with updated Moderna COVID-19 vaccination versus no updated vaccination, by underlying medical condition

Across all five high-risk populations, vaccination with the updated Moderna COVID-19 vaccine also resulted in cost savings from a healthcare payer perspective and from a societal perspective (considering productivity losses due to vaccine administration, adverse events, outpatient care, hospitalization, hospitalization recovery and severe long COVID), both at the population (**Fig. 3a**) and patient level (**Fig. 3b**). At the population level, the largest savings were among CLD, DM and CVD populations, as these conditions are more prevalent. The updated vaccination provided both cost savings and QALYs saved, thus, the ICER was dominant for all high-risk groups (**Table S9**). In the scenario analysis assessing two-dose vaccination (in fall and spring) in high-risk groups aged ≥65 years, results remained cost-saving across all conditions. At the patient level, vaccination also resulted in overall healthcare and societal cost savings per patient, with the largest cost savings for DM, IC, and CKD patients, as these groups have the largest treatment costs. The return on investment (ROI) was positive in all high-risk groups, with the highest ROI seen for CKD patients, i.e., a return of $2.60 (healthcare payer perspective) or $2.80 (societal perspective) for every $1 invested (**Fig. 3b**) (see **Supplemental File S3** for societal perspective results).

**Fig 3.**
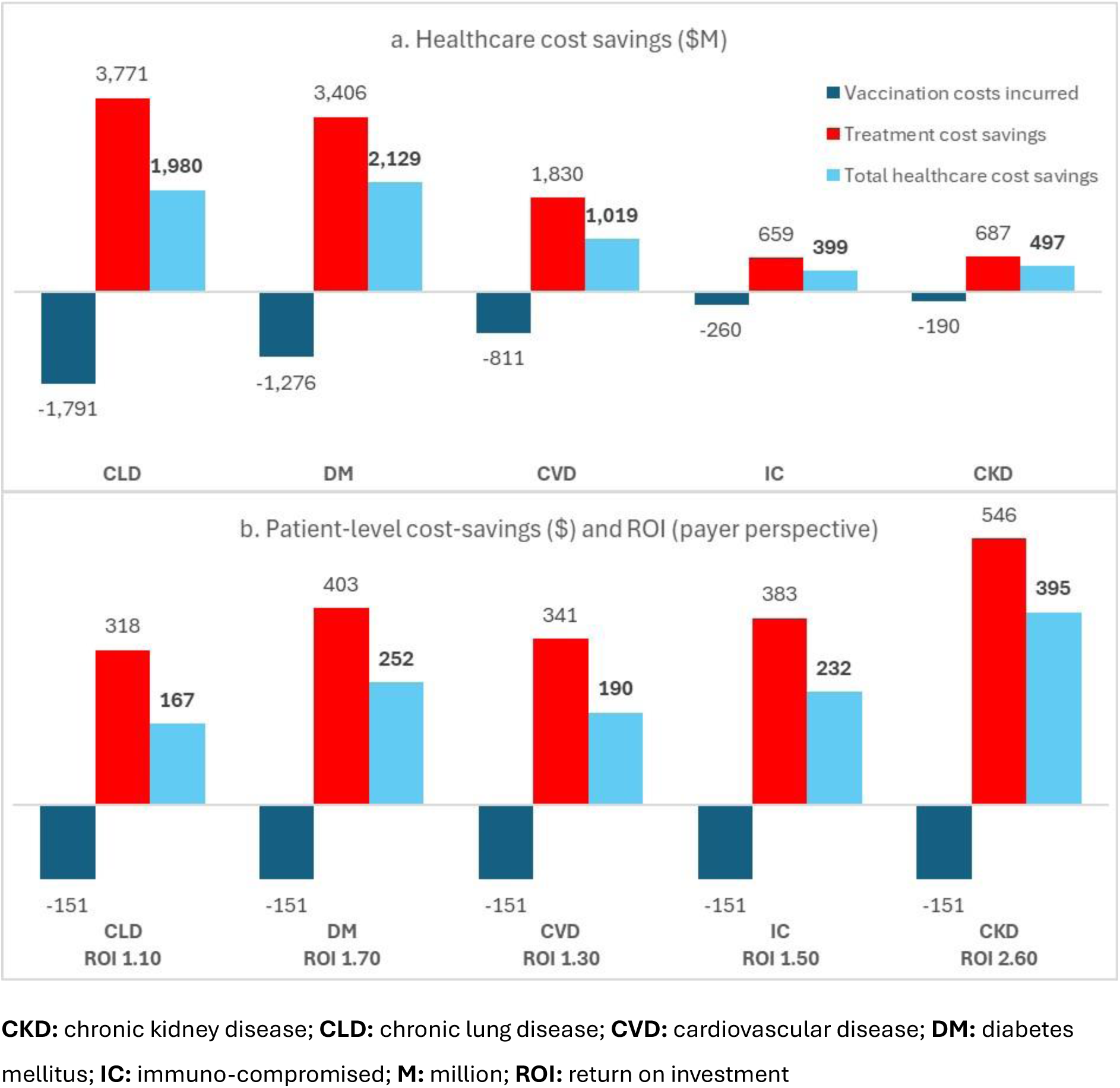
**a)** Total healthcare cost savings ($M), and b) patient-level cost savings ($) and ROI (payer perspective) with updated Moderna COVID-19 vaccination versus no updated vaccination

In all scenario analyses, updated Moderna COVID-19 vaccination remained dominant (i.e., providing more health benefits and cost savings) versus no vaccination. The impact of scenario analyses on healthcare cost savings and on QALYs saved was comparable for all high-risk populations, therefore, the results for CLD (which had the largest costs) are presented in **Fig 4** (see **Supplemental File S2** for results in other high-risk groups). Scenario analyses with the largest impact on healthcare cost savings were: an increased rate of COVID-19 hospitalization (increasing base case savings by $2,065.57M); varying the incidence of COVID-19 (i.e., a higher incidence would increase base case savings by $863.90M and a lower incidence would reduce base case savings by $971.80M); and assuming an alternative post-infection cost (reducing base case savings by $1,017.72M) (**Fig. 4**). Similarly, the scenario analyses with the largest impact on health benefits were: an increased rate of hospitalization (resulting in 135.46K more QALYs saved than in the base case); and varying the incidence of COVID-19 (a higher incidence would increase base case QALYs saved by 44.20K, and a lower incidence would decrease them by 47.79K) (**Fig. 4**).

**Fig 4.**
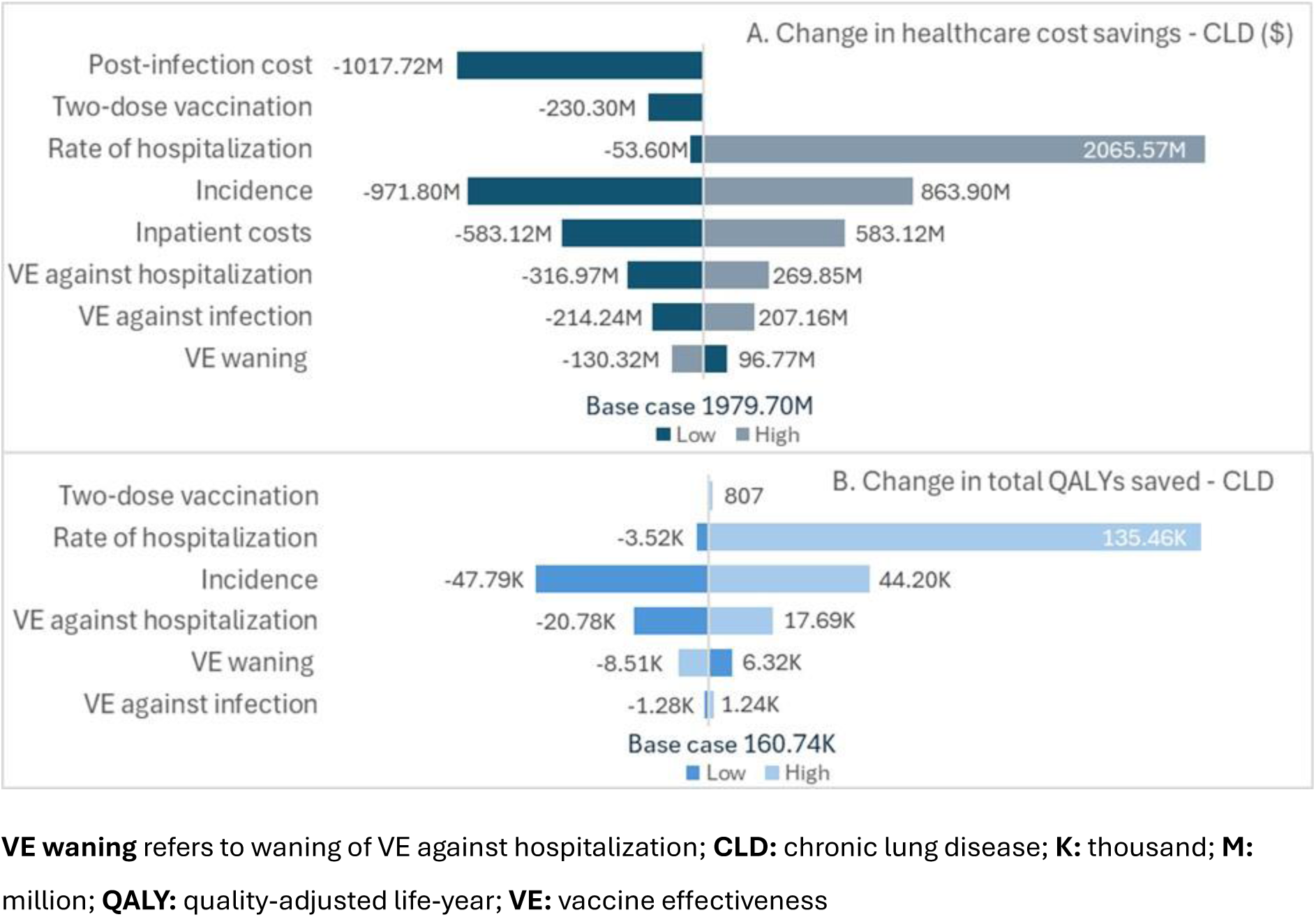
Scenario analyses: Tornado diagram of changes versus base case for a) Healthcare cost savings and b) QALYs saved, among CLD patients (healthcare payer perspective)

With an increased VCR of 70%, the updated Moderna vaccination could avert an additional 16,488 (CKD) to 165,352 (DM) hospitalizations (**Fig. 5a**) and an additional 2,061 (CKD) to 19,634 (DM) COVID-19 deaths compared with the current VCR among high-risk populations (**Fig. 5b**). An increase in the current VCR of 10% could also avert an additional 10% of hospitalizations and deaths in high-risk populations compared with the base case (**Fig. 5**), resulting in healthcare cost savings of $2,178M, increasing to $4,697M for a VCR of 70% for CLD alone.

**Fig 5.**
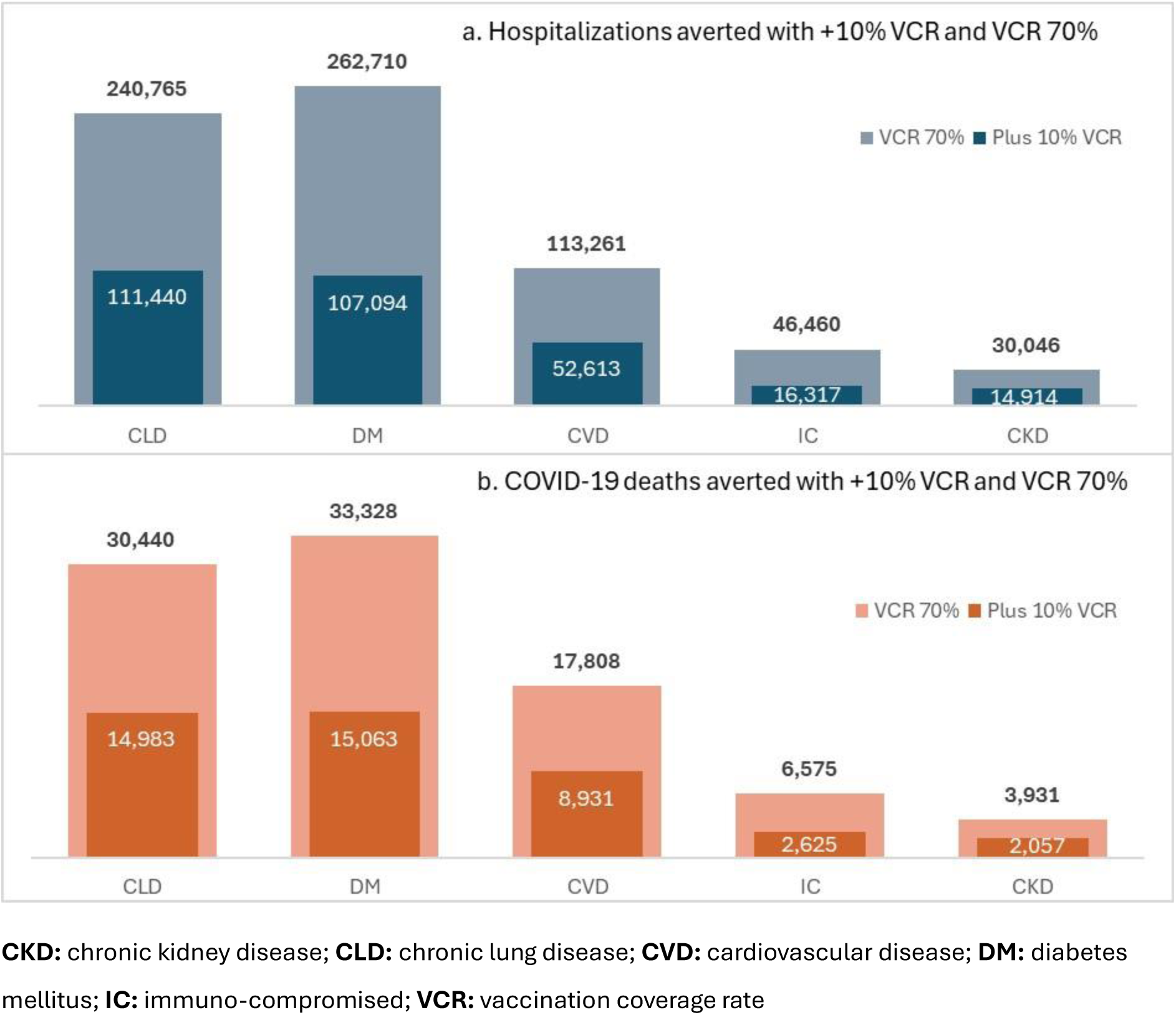
Impact of increasing current VCR (by 10% and to 70% VCR) on a) COVID-19 hospitalizations averted, and b) deaths averted with updated Moderna COVID-19 vaccination versus no updated vaccination

Healthcare cost savings increased for VCR plus 10% and for 70% VCR, respectively, in all high-risk populations (i.e., DM: $2,342M and $5,707M; CVD: $1,121M and $2,431M; IC: $439M and $1,229M; CKD: $546M and $1,096M).

### Secondary analysis: Moderna versus Pfizer/BioNTech updated COVID-19 vaccination

In the secondary analysis, vaccination with Moderna’s updated COVID-19 vaccine was compared with Pfizer/BioNTech’s updated COVID-19 vaccine during the 2023/24 season. The model estimated that more hospitalizations and COVID-19 deaths would be averted in all high-risk populations with the Moderna vaccine. Specifically, an additional 942 (CKD) to 13,105 (DM) hospitalizations would be averted (**Fig 6a**), and additional 130 (CKD) to 1,844 (DM) COVID-19 deaths would be averted (**Fig 6b**) with vaccination with Moderna’s vaccine versus Pfizer/BioNTech’s vaccine during Fall/Winter 23/24.

**Fig 6.**
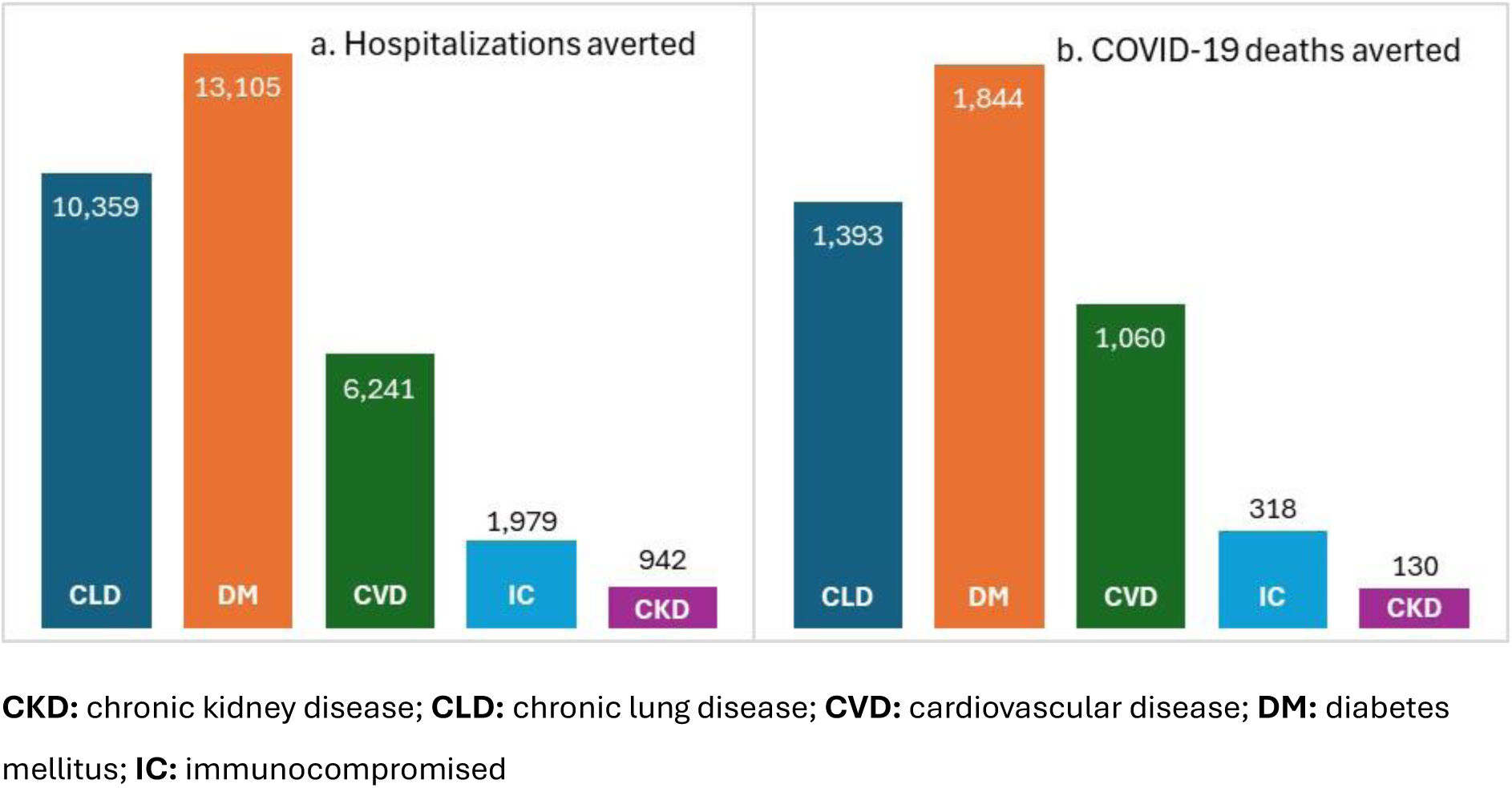
**a)** COVID-19 hospitalizations averted and b) deaths averted with Moderna versus Pfizer/BioNTech updated COVID-19 vaccination

In addition, vaccination with Moderna’s versus Pfizer/BioNTech’s updated COVID-19 vaccine produced cost savings from the healthcare payer perspective and societal perspective, making it the dominant strategy. From a healthcare payer perspective, cost savings were: $370M for CLD, $434M for DM, $303M for CVD, $90M for IC, and $74M for CKD, and from the societal perspective: $404M for CLD, $492 for DM, $331 for CVD, $99M for IC, and $78M for CKD. At the patient level, vaccination with Moderna’s updated vaccine versus Pfizer/BioNTech’s updated vaccine resulted in healthcare cost savings of $31-59 per patient and with societal cost savings of $34-62 per patient, depending on high-risk population. Full results and sensitivity analyses are presented in **Supplemental File S4**.

## 4. Discussion

In this analysis, vaccination of high-risk populations in the US with the updated Moderna COVID-19 vaccine during the 2023/24 season was estimated to provide significant health gains in terms of prevention of symptomatic cases, hospitalizations, deaths, and long COVID cases, and was cost-saving. Specifically, each dose of the updated Moderna COVID-19 vaccine administered to individuals with underlying medical conditions associated with severe COVID-19 resulted in cost savings from both the healthcare payer and societal perspectives. The estimated ROI was high i.e., every $1 invested in the updated Moderna COVID-19 vaccine returned $1.10–$2.60 from the healthcare payer perspective and $1.30–$2.80 from the societal perspective (including savings in productivity costs). Vaccination was, therefore, the dominant strategy across all high-risk populations assessed. Increasing the current VCR by 10% or to 70% VCR provided even greater protection in these at-risk populations while saving more healthcare and societal costs e.g., healthcare cost savings with the current VCR were: $1,980M (increasing to $2,178M–$4,697M, for plus 10% VCR to 70% VCR, respectively) for CLD; $2,129M (increasing to $2,342M–$5,707M) for DM; $1,019M (increasing to $1,121M–$2,431M) for CVD; $399M (increasing to $439M–$1,229M) for IC; and $497M (increasing to $546M–$1,096M) for CKD. When compared with the updated Pfizer/BioNTech’s COVID-19 vaccine, Moderna’s vaccine provided more health gains while being cost-saving (i.e., dominant strategy), based on its higher VE observed in real-world studies [17, 19]. At the patient level, healthcare cost savings per patient vaccinated with the updated Moderna versus Pfizer/BioNTech’s vaccine were $31–$59, depending on the high-risk population. Overall, this analysis has shown that annual COVID-19 vaccination during the post-pandemic era results in significant clinical and financial benefits at the patient and system level. These benefits increase significantly with an increase in VCR.

Moreover, the difference between the mRNA vaccines, in terms of protection in high-risk populations, may lead to a significant difference in societal and financial benefits at the system and individual levels, and thus, be relevant for decision making about choice of vaccine type.

The risks of severe COVID-19 outcomes, such as hospitalization and death, are higher in people with underlying medical conditions, and increase with multiple underlying conditions [53, 54]. For instance, the risk of hospitalization was over six times higher among working-age adults with CLD versus people without underlying conditions [54], and the risk of death was 1.5 (95% CI 1.4–1.7) times higher with the presence of one underlying condition rising to 3.8 (95% CI 3.5– 4.2) times higher with over 10 underlying conditions in the US [53]. This analysis assessed outcomes per medical condition, and thus, may underestimate the clinical and financial benefits of vaccination in individuals suffering from multiple conditions. This analysis also assessed the incremental benefits of annual COVID-19 vaccination, regardless of vaccination history. The low adult vaccination rate in the US presents a large unmet need, with particularly important consequences – both clinical and financial, in high-risk groups. Thus, it remains critical for public health authorities, healthcare systems, payers, and healthcare providers to increase VCR and reduce the burden of COVID-19. Key strategies identified to support this goal include facilitating co-administration of influenza and COVID-19 vaccination, integrating COVID-19 vaccination into routine care delivery e.g., to improve equitable access, and supporting healthcare providers in addressing patient barriers and providing vaccination [54].

The latest guidance from the CDC (October 2024) recommends two doses of COVID-19 vaccination six months apart in people at increased risk of severe outcomes (moderately to severely immunocompromised) as well as those aged 65 years and older [55]. Our analysis results showed two-dose vaccination in high-risk groups was cost-saving, thus reinforcing the importance of increasing vaccination rates for both the first dose in the fall and also the second dose following later in the season in these populations.

Our model results were in line with other economic analyses in high-risk populations, including IC adults in Canada [16] and France [56], where updated Moderna COVID-19 Fall 2023 vaccination provided more health gains at a cost-saving, and was, therefore, the dominant strategy versus updated Pfizer/BioNTech vaccination. The model results in high-risk populations were also in line with the previous economic analysis in the US adult general population, showing that updated Moderna COVID-19 vaccination was highly cost-effective versus no updated vaccination and versus Pfizer/BioNTech updated vaccination, from a healthcare payer perspective [30]. From a societal perspective, in adults aged ≥65 years who are at higher risk of severe outcomes, updated Moderna COVID-19 vaccination was cost-saving versus no updated vaccination and versus Pfizer/BioNTech updated vaccination [30]. Similarly, a recent US economic analysis in the general population aged ≥65 years estimated that a one-dose updated mRNA COVID-19 Fall vaccination was highly cost-effective versus no updated vaccination [57].

Our economic analysis is subject to limitations. There is large uncertainty related to the epidemiology of COVID-19, as new variants continue to emerge. As such, it is difficult to predict the incidence, disease severity, and effectiveness of current and future vaccines against new variants. In addition, future vaccine uptake rates in the high-risk population are not known.

Scenario analyses were conducted to assess the impact of uncertainty around key input parameters and confirmed that the findings were robust to variations in these inputs. In the absence of high-risk population data, the model relied on the US general population data [30] for several inputs, including COVID-19 incidence, probability of post-discharge COVID-19 death, proportions with post COVID conditions, long COVID and severe long COVID, utility estimates, and outpatient costs. The use of general population data for high-risk groups is likely to result in conservative estimates, for example, data suggest that IC patients are at increased risk of long COVID (OR 1.5 [58]), although uncertainty exists on the distinction between long COVID and persistent COVID-19 infections with prolonged viral shedding in IC patients [59, 60]. Data informing post-discharge COVID-19 deaths, however, were based on a study that included 86% of patients with high-risk conditions [61]. The risks of hospitalization may also be conservative estimates, as they were considered for each high-risk population separately, however, in reality people are likely to have multiple high-risk conditions, thus may be at higher risk of hospitalization or death. The hospitalization risk in IC patients (RR 1.20) [34] was lower than estimates reported in the literature, e.g., ranging from 1.3 to 13.1 [62], possibly due to the restrictive IC definition used [34]. Data on productivity loss due to long COVID in high-risk populations are limited and subject to uncertainty, as symptoms associated with long COVID may potentially overlap with exacerbations of underlying conditions triggered by long COVID [63]. Available data indicate that long COVID is associated with considerable productivity loss [64], and is an independent risk factor for acquiring disability [64–66]. Taking a conservative approach, the model productivity loss due to long COVID was only applied to severe long COVID cases, with the number of days lost estimated using the most relevant available sources [66, 67]. This analysis was conducted using a static model, which only accounts for direct vaccination protection in vaccinated individuals. Thus, it conservatively does not account for indirect protection in unvaccinated people, due to decreased transmission in the population [30, 68]. The analysis assumed that the updated vaccines were well-matched to circulating variants, and results are, therefore, also expected to apply to future well-matched versions of Moderna’s updated COVID-19 vaccine, expected to be available for the following seasons from 2024 onwards. For the comparison to Pfizer/BioNTech’s vaccine, relative VE data were based on a single study available at the time of analysis based on the bivalent original and BA4/5 containing mRNA COVID-19 vaccines [17]. However, recent data from a meta-analysis support the assumptions made [19]. Finally, the CDC published estimates of the VE of updated COVID-19 Fall 2023 vaccines against symptomatic infection from testing during September 2023 to January 2024, reporting a VE of 54% (95% CI 46–60%) versus no updated vaccination, with data available up to 119 days after vaccination [3]. These estimates are higher than the base case VE against infection (34.50%) used in the model, as such, the benefits of vaccination could be expected to be even greater than those estimated in this analysis.

### 4.1. Conclusion

COVID-19 continues to place a burden on healthcare systems and patients. As vaccination coverage remains low, there is an unmet need, especially in populations with underlying medical conditions associated with severe COVID-19 outcomes. An annual vaccination with updated vaccines that target the circulating variants is, therefore, recommended for everyone aged ≥6 months. This analysis estimated that vaccination with the updated 2023/24 Moderna COVID-19 vaccine in high-risk populations (including CLD, CKD, CVD, DM, and IC) provided significant clinical benefits and cost savings, which could be substantially improved with even a modest 10% increase in vaccination coverage. Vaccination was cost-saving and provided a positive return on investment of up to $2.60 per $1 invested in vaccination, which could help to mitigate the high healthcare costs in these high-risk groups even further. Overall, compared with no updated vaccination, and compared with Pfizer/BioNTech’s updated vaccine, the updated Moderna COVID-19 vaccine provided protection that resulted in fewer clinical cases, and was cost-saving at the individual and system levels in high-risk populations.

## Data Availability

The data presented in this study are available upon reasonable request from the corresponding author.

## 5.2 Compliance with Ethics Guidelines

This article is based on previously conducted studies and does not contain any new studies with human participants or animals performed by any of the authors.

## 5.3 Medical writing, Editorial, and other Assistance

Medical writing and editorial assistance were provided by Kavi Littlewood (Littlewood Writing Solutions) in accordance with Good Publication Practice (GPP 2022) guidelines, funded by Moderna, Inc., and under the direction of the authors.

## 5.5 Author contributions

All authors contributed to the study conception and design. Material preparation, data collection and analysis were performed by Keya Joshi, Ekkehard Beck, Mariia Dronova, and Ewelina Paterak. The first draft of the manuscript was written by Mariia Dronova and Ewelina Paterak and all authors commented on previous versions of the manuscript. All authors read and approved the final manuscript.

