## Supplemental material for "Clinical impact and cost-effectiveness of updated 2023/24 COVID-19 mRNA vaccination in high-risk populations in the United States"

***Contents:***

**File S1. Methods tables**

**File S2. Indirect costs and QALYs**

**File S3. Primary analysis results**

**File S4. Secondary analysis results**

**File S1. Methods tables**

**Table S1. Population size by condition and age group**

| Age, y | CLD | DM | CVD | IC | CKD |
|---|---|---|---|---|---|
| 18-29 | 1,520,407 | 1,617,426 | 370,602 | 729,647 | 114,131 |
| 30-39 | 1,285,417 | 1,367,440 | 546,976 | 616,874 | 96,491 |
| 40-49 | 1,182,300 | 3,653,659 | 1,003,115 | 1,006,191 | 88,751 |
| 50-64 | 11,904,738 | 9,376,977 | 4,669,876 | 2,922,188 | 885,617 |
| 65-74 | 12,573,728 | 7,920,487 | 4,641,925 | 1,178,233 | 1,497,944 |
| 75-84 | 6,121,910 | 3,856,335 | 3,824,726 | 442,810 | 729,320 |
| ≥85 | 2,415,331 | 1,521,474 | 1,509,003 | 139,998 | 287,745 |
| **Total** | **37,003,832** | **29,313,799** | **16,566,223** | **7,035,940** | **3,700,000** |

**CKD:** chronic kidney disease; **CLD:** chronic lung disease; **CVD:** cardiovascular disease; **DM:** diabetes mellitus; **IC:** immune-compromised; **y:** year

**Table S2. COVID-19 predicted incidence rates 2023-2024 (general population)**

| Age group | Sep 23 | Oct 23 | Nov 23 | Dec 23 | Jan 24 | Feb 24 | Mar 24 | Apr 24 | May 24 | Jun 24 | Jul 24 | Aug 24 |
|---|---|---|---|---|---|---|---|---|---|---|---|---|
| **Base case incidence rates** | | | | | | | | | | | | |
| 18-29y | 1.88 | 2.56 | 3.05 | 3.40 | 3.11 | 2.31 | 1.80 | 1.22 | 0.88 | 0.62 | 0.50 | 0.43 |
| 30-39y | 1.88 | 2.57 | 3.08 | 3.46 | 3.18 | 2.37 | 1.84 | 1.24 | 0.89 | 0.62 | 0.49 | 0.42 |
| 40-49y | 1.82 | 2.49 | 3.00 | 3.38 | 3.12 | 2.34 | 1.82 | 1.22 | 0.88 | 0.61 | 0.48 | 0.41 |
| 50-64y | 1.45 | 2.01 | 2.47 | 2.86 | 2.71 | 2.07 | 1.62 | 1.08 | 0.77 | 0.52 | 0.40 | 0.33 |
| 65-74y | 0.86 | 1.22 | 1.53 | 1.84 | 1.82 | 1.43 | 1.14 | 0.76 | 0.53 | 0.35 | 0.27 | 0.22 |
| 75-84y | 0.62 | 0.89 | 1.13 | 1.36 | 1.36 | 1.08 | 0.86 | 0.58 | 0.40 | 0.27 | 0.20 | 0.16 |
| ≥85y | 0.58 | 0.82 | 1.04 | 1.25 | 1.25 | 0.99 | 0.79 | 0.53 | 0.37 | 0.24 | 0.18 | 0.15 |
| **Scenario: Low incidence** | | | | | | | | | | | | |
| 18-29y | 1.85 | 2.19 | 2.14 | 1.95 | 1.54 | 1.08 | 0.84 | 0.58 | 0.43 | 0.30 | 0.23 | 0.18 |
| 30-39y | 1.91 | 2.27 | 2.21 | 2.02 | 1.60 | 1.12 | 0.87 | 0.60 | 0.43 | 0.30 | 0.23 | 0.18 |
| 40-49y | 1.88 | 2.25 | 2.22 | 2.03 | 1.62 | 1.14 | 0.88 | 0.60 | 0.44 | 0.30 | 0.23 | 0.18 |
| 50-64y | 1.67 | 2.06 | 2.08 | 1.96 | 1.59 | 1.13 | 0.87 | 0.59 | 0.42 | 0.28 | 0.21 | 0.16 |
| 65-74y | 1.15 | 1.47 | 1.54 | 1.51 | 1.27 | 0.91 | 0.71 | 0.48 | 0.33 | 0.22 | 0.16 | 0.12 |
| 75-84y | 0.87 | 1.12 | 1.19 | 1.18 | 1.00 | 0.72 | 0.56 | 0.38 | 0.27 | 0.18 | 0.13 | 0.10 |
| ≥85y | 0.80 | 1.03 | 1.09 | 1.08 | 0.91 | 0.66 | 0.51 | 0.35 | 0.24 | 0.16 | 0.12 | 0.09 |
| **Scenario: High incidence** | | | | | | | | | | | | |
| 18-29y | 2.65 | 3.76 | 4.62 | 5.19 | 4.70 | 3.48 | 2.74 | 1.92 | 1.48 | 1.12 | 0.98 | 0.93 |
| 30-39y | 2.57 | 3.68 | 4.57 | 5.20 | 4.76 | 3.53 | 2.78 | 1.94 | 1.48 | 1.11 | 0.96 | 0.90 |
| 40-49y | 2.44 | 3.49 | 4.35 | 4.97 | 4.57 | 3.41 | 2.68 | 1.87 | 1.42 | 1.07 | 0.92 | 0.86 |
| 50-64y | 1.76 | 2.56 | 3.26 | 3.83 | 3.62 | 2.75 | 2.18 | 1.50 | 1.13 | 0.83 | 0.70 | 0.64 |
| 65-74y | 0.90 | 1.33 | 1.74 | 2.12 | 2.09 | 1.64 | 1.31 | 0.90 | 0.66 | 0.48 | 0.39 | 0.35 |
| 75-84y | 0.63 | 0.93 | 1.22 | 1.50 | 1.49 | 1.17 | 0.94 | 0.65 | 0.48 | 0.34 | 0.28 | 0.25 |
| ≥85y | 0.58 | 0.86 | 1.13 | 1.38 | 1.37 | 1.07 | 0.86 | 0.59 | 0.44 | 0.31 | 0.26 | 0.23 |

**y:** year

The rate of hospitalization given symptomatic COVID-19 infection in the unvaccinated general population (10.54% for adults) was based on claims data [1] and adjusted for high-risk groups by applying a risk ratio (RR) based on hazard ratios (HRs) from data on disease burden in patients with medical conditions [2]. The RRs were estimated as an optimal minimax transformation of HRs [3] (**Table S3**).

For CKD, CLD, CVD and DM patients, the distribution by in-hospital level of care (no ICU or ventilator; ICU only; ventilator) was derived from the CDC COVID-NET hospitalizations tracker in the season 2023-2024 [4]. For patients aged 65 years and older, these estimates were then redistributed by the model age groups, using data provided in CDC Morbidity and Mortality Weekly Report (MMWR) [5]. Due to the lack of condition-specific data, the general population estimates were assumed to be applicable for these populations. The distribution for the IC population was estimated by applying a RR to general population CDC data [4, 5]. The RR for ICU admission (RR of 1.30) was derived by converting the reported OR of 1.40 [6]. The same RR was assumed for the increased risk of ventilator use in IC patients (**Table S3**).

For CVD, DM and IC patients, in-hospital mortality was estimated by applying an RR to the general population mortality due to COVID-19 [4]: RR of 1.23 for DM [7]; RR of 1.62 for CVD [8] ; and RR of 1.74 for IC [6]. For CKD and CLD patients, in-hospital mortality rates were informed by data reported for the general population [4], in line with previous studies [9, 10]. In-hospital mortality for the general population was derived from the CDC COVID-NET hospitalizations tracker in the season 2023-2024 [4]. For patients 65 years and older, these estimates were then redistributed by the model age groups, using CDC data [5] (**Table S3**).

The risk of hospital re-admission by location of care was estimated by applying a RR to general population data [11, 12], with RRs based on converting ORs (**Table S3**).

The risk of post discharge mortality was based on previous estimates from a US economic analysis of COVID-19 [12], due to the lack of data in high-risk patients, and assumed to be the same for ICU only and ventilator (**Table S3**).

Following symptomatic infection, there is an age-dependent risk of myocarditis. A portion of infections are treated in hospital, and the risk of this more severe type of COVID-19 increases with age. Data were from the general population (**Table S4**).

**Table S3. Hospitalization rates by high-risk population**

| | Base case | Scenario: LB for hospitalization rate | Scenario: UB for hospitalization rate |
|---|---|---|---|
| **Hospitalization rates, given infection (%)** | | | |
| All ages, CLD | 14.90% | 14.56% | 28.06% |

| | | | | | |
|---|---|---|---|---|---|
| All ages, DM | 19.05% | 18.70% | 28.82% | | |
| All ages, CVD | 16.19% | 15.72% | 33.46% | | |
| All ages, IC | 12.58% | 12.13% | 29.31% | | |
| All ages, CKD | 19.8% | 18.5% | 49.4% | | |
| **In-hospital level of care, by age (%) – other reports (general population)** | | | | | |
| | **No ICU or ventilator** | **ICU only** | **Ventilator** | | |
| 18-49y | 80.04% | 14.89% | 5.07% | | |
| 50-64y | 71.74% | 18.83% | 9.43% | | |
| 65-74y | 68.24% | 21.50% | 10.26% | | |
| 75-84y | 84.50% | 8.86% | 6.64% | | |
| ≥85y | 74.84% | 17.50% | 7.66% | | |
| **In-hospital level of care, by age (%) – IC report** | | | | | |
| | **No ICU or ventilator** | **ICU only** | **Ventilator** | | |
| 18-49y | 74.24% | 19.21% | 6.54% | | |
| 50-64y | 63.53% | 24.30% | 12.17% | | |
| 65-74y | 59.02% | 27.74% | 13.24% | | |
| 75-84y | 80.00% | 11.43% | 8.57% | | |
| 85+y | 67.53% | 22.59% | 9.88% | | |
| **Age group** | **In-hospital outcomes** | | | | |
| **In-hospital mortality, by age (%), any level of care,** | | | | | |
| | **CLD (=general population)** | **DM** | **CVD** | **IC** | **CKD** |
| 18-49y | 1.34% | 1.61% | 2.09% | 2.28% | 2.10% |
| 50-64y | 5.86% | 7.04% | 9.14% | 9.98% | 4.00% |
| 65-74y | 9.35% | 11.23% | 14.59% | 15.92% | 6.40% |
| 75-84y | 7.25% | 8.70% | 11.31% | 12.34% | 4.90% |
| ≥85y | 7.08% | 8.51% | 11.05% | 12.06% | 4.10% |
| **Hospital re-admission rate, by location of care** | | | | | |
| | **No ICU or ventilator** | **ICU only** | **Ventilator** | | |
| All ages, CLD | 4.03% | 3.86% | 2.09% | | |
| All ages, DM | 4.65% | 4.46% | 2.41% | | |
| All ages, CVD | 5.08% | 4.87% | 2.63% | | |
| All ages, IC | 4.31% | 4.13% | 2.24% | | |
| All ages, CKD | 8.70% | 8.70% | 8.70% | | |
| **Post discharge mortality, by location of care (%), general population** | | | | | |
| | **No ICU or ventilator** | **ICU only** | **Ventilator** | | |
| All ages | 5.36% | 10.40% | 10.40% | | |

**CKD:** chronic kidney disease; **CLD:** chronic lung disease; **CVD:** cardiovascular disease; **DM:** diabetes mellitus; **IC:** immune-compromised; **LB/UB:** lower bound/ upper bound; **y:** year

**Table S4. Long COVID and myocarditis (infection-related), general population (and CKD population)**

| Age group | Proportion with any long COVID | Proportion with severe long COVID (of those with long COVID) | % experiencing myocarditis (infection-related) |
|---|---|---|---|
| 18-29y | 26.90% | 6.9% (13.9% CKD) | 0.08% |
| 30-39y | 31.70% | 6.9% (13.9% CKD) | 0.07% |
| 40-49y | 32.60% | 6.9% (13.9% CKD) | 0.09% |
| 50-64y | 32.30% | 6.9% (13.9% CKD) | 0.14% |
| 65-74y | 28.00% | 6.9% (13.9% CKD) | 0.16% |
| 75-84y | 22.70% | 6.9% (13.9% CKD) | 0.21% |
| ≥85y | 18.50% | 6.9% (13.9% CKD) | 0.21% |

**y:** year

**Table S5. Direct healthcare costs and indirect costs**

| Model parameter | Base case | Scenario: LB hospitalization cost | Scenario: UB hospitalization cost | Scenario: Alternative post-infection cost |
|---|---|---|---|---|
| **Vaccination costs** | | | | |
| Spikevax vaccine cost* | $129.50 | base case | base case | base case |
| Comirnaty vaccine cost* | $129.00 | base case | base case | base case |
| Administration cost* | $20.33 | base case | base case | base case |
| **Short-term infection: not hospitalized** | | | | |
| Proportion not hospitalized who seek outpatient care* | 41.00% | base case | base case | base case |
| Cost of acute outpatient care* | $460.57 | base case | base case | base case |
| **Short-term infection: hospitalized** | | | | |
| No ICU or ventilation * | $15,089.00 | $11,316.75 | $18,861.25 | base case |
| ICU only * | $27,058.00 | $20,293.50 | $33,822.50 | base case |
| Ventilator * | $71,367.00 | $53,525.25 | $89,208.75 | base case |
| Cost of inpatient follow-up (per case)* | $1,075.22 | base case | base case | base case |
| DM -No ICU or ventilation * | $16,150.78 | $12,113.08 | $20,188.47 | base case |
| DM - ICU only * | $28,962.01 | $21,721.50 | $36,202.51 | base case |
| DM - Ventilator * | $76,388.92 | $57,291.69 | $95,486.15 | base case |
| CVD -No ICU or ventilation * | $16,632.48 | $12,474.36 | $20,790.60 | base case |
| CVD - ICU only * | $29,825.81 | $22,369.36 | $37,282.26 | base case |
| CVD - Ventilator * | $78,667.26 | $59,000.44 | $98,334.07 | base case |
| IC -No ICU or ventilation * | $18,734.79 | $14,051.09 | $23,418.49 | base case |
| IC - ICU only * | $33,595.73 | $25,196.80 | $41,994.66 | base case |
| IC - Ventilator * | $88,610.63 | $66,457.97 | $110,763.29 | base case |
| CKD -No ICU or ventilation * | $24,572.75 | $18,429.56 | $30,715.94 | base case |
| CKD - ICU only * | $44,064.52 | $33,048.39 | $55,080.65 | base case |
| CKD - Ventilator * | $116,222.65 | $87,166.98 | $145,278.31 | base case |
| **Other costs** | | | | |
| Post-infection, not hospitalized (18-64 years)*** | $3,262.00 | base case | base case | $504.00 |
| Post-infection, not hospitalized (≥ 65 years)*** | $6,930.00 | base case | base case | $1,666.00 |
| Post-infection, hospitalized (18-64 years)*** | $3,262.00 | base case | base case | $504.00 |
| Post-infection, hospitalized (≥ 65 years)*** | $6,930.00 | base case | base case | $1,666.00 |
| Myocarditis, infection-related* | $14,130.00 | base case | base case | base case |
| Adverse events, grade 3 local (all ages)* | $7.59 | base case | base case | base case |
| Adverse events, grade 3 systemic (all ages)* | $11.49 | base case | base case | base case |
| Adverse events, anaphylaxis (all ages)* | $3,412.81 | base case | base case | base case |
| Adverse events, myocarditis / Pericarditis (applies to ages 18-39 only)* | $3,584.48 | base case | base case | base case |

| Indirect costs - Model parameter | | | | | Value |
|---|---|---|---|---|---|
| **Labor force** | | | | | |
| | **CLD** | **DM** | **CVD** | **IC** | **CKD** |
| 18-29y | 42.51% | 55.50% | 52.34% | 36.39% | 69.8% |
| 30-39y | 49.20% | 64.23% | 60.58% | 42.11% | 69.8% |
| 40-49y | 48.91% | 63.86% | 60.22% | 41.87% | 69.8% |
| 50-64y | 41.27% | 53.88% | 50.82% | 35.33% | 69.8% |
| 65-74y | 15.72% | 20.52% | 19.36% | 13.46% | 0.0% |
| ≥75y | 4.83% | 6.31% | 5.95% | 4.14% | 0.0% |
| Daily wage* | | | | | $262.93 |
| **Productivity loss (days)** | | | | | |
| Time loss for vaccine administration* | | | | | 0.04 |
| Short-term infection: not hospitalized* | | | | | 3.57 |
| Short-term infection: hospitalized, No ICU or ventilator* | | | | | 7.50 |
| Short-term infection: hospitalized, ICU only* | | | | | 7.50 |
| Short-term infection: hospitalized, Ventilator* | | | | | 19.75 |
| IC - Short-term infection: hospitalized, No ICU or ventilator** | | | | | 11.2 |
| IC - Short-term infection: hospitalized, ICU only** | | | | | 19.9 |
| IC - Short-term infection: hospitalized, Ventilator** | | | | | 19.9 |
| CKD - Short-term infection: hospitalized, No ICU or ventilator | | | | | 5.43 |
| CKD - Short-term infection: hospitalized, ICU only | | | | | 8.55 |
| CKD - Short-term infection: hospitalized, Ventilator | | | | | 15.05 |
| Hospitalization recovery days lost* | | | | | 33.43 |
| DM - Post-infection: Additional severe long COVID days lost* | | | | | 33 |
| CLD - Post-infection: Additional severe long COVID days lost | | | | | 66 |
| CVD - Post-infection: Additional severe long COVID days lost | | | | | 85 |
| IC - Post-infection: Additional severe long COVID days lost | | | | | 42 |
| CKD - Post-infection: Additional severe long COVID days lost | | | | | 54.89 |

* General population; *** High-risk population; **CKD:** chronic kidney disease; **CLD:** chronic lung disease; **CVD:** cardiovascular disease; **DM:** diabetes mellitus; **IC:** immune-compromised; **LB/UB:** lower bound/ upper bound; **y:** year

**Table S6. Utility inputs, general population**

| Model parameter | Value |
|---|---|
| **Baseline utility** | |
| 18-29 years | 0.9205 |
| 30-39 years | 0.9055 |
| 40-49 years | 0.8750 |
| 50-64 years | 0.8490 |
| 65-74 years | 0.8255 |
| 75-84 years | 0.7865 |
| ≥85 years | 0.7530 |
| **Utility decrements** | |
| Symptomatic infection, not hospitalized (all ages) * | 0.006 |
| Hospitalized, No ICU or ventilator (all ages) | 0.027 |
| Hospitalized, ICU only (all ages) | 0.027 |
| Hospitalized, ICU with ventilator (all ages) | 0.054 |
| Hospital readmission | 0.011 |
| Post-infection, not hospitalized (all ages) | 0.0277 |
| Post-infection, hospitalized (all ages) | 0.1224 |
| Myocarditis, infection-related (all ages) | 0.0019 |
| Adverse events, grade 3 local (all ages) | 0.0003 |

| | |
|---|---|
| Adverse events, grade 3 systemic (all ages) | 0.0011 |
| Adverse events, anaphylaxis (all ages) | 0.0019 |
| Adverse events, myocarditis / Pericarditis (applies to ages 18-39 only) | 0.0019 |

**ICU**: Intensive care unit, **QALY**: quality-adjusted life-year; *Regardless of medical attendance

**Table S7. Vaccine effectiveness of updated Moderna and updated Pfizer/BioNTech COVID-19 vaccines in high-risk populations**

| **Moderna - Infection** | | **Moderna - Hospitalization** | |
|---|---|---|---|
| **Initial VE** | **Waning** | **Initial VE** | **Waning** |
| Base case: 34.50% | 4.75% | 58.70% | 1.37% |
| Scenario VE against infection: | | | |
| 31.20% –37.60% | base case | base case | base case |
| Scenario VE against hospitalization: | | | |
| base case | base case | 51.30%–65.00% | base case |
| Scenario waning of VE against hospitalization: | | | |
| base case | base case | base case | 0.62%–2.38% |
| **Pfizer/BioNTech - Infection** | | **Pfizer/BioNTech - Hospitalization** | |
| **Initial VE** | | **Initial VE** | |
| **CLD - Base case VE** (rVE against infection 3.10%; rVE against hospitalization 11.90%) | | | |
| 32.40% (scenario 30.54–34.24%) | | 53.10% (scenario 49.51–56.48%) | |
| **DM - Base case VE** (rVE against infection 3.70%; rVE against hospitalization 15.10%) | | | |
| 31.98% (scenario 30.17–33.70%) | | 51.35% (scenario 47.72–54.76%) | |
| **CVD - Base case VE** (rVE against infection 7.40%; rVE against hospitalization 14.70%) | | | |
| 29.27% (scenario 27.38–31.13%) | | 51.58% (scenario 48.31–54.62%) | |
| **IC - Base case VE** (rVE against infection 4.80%; rVE against hospitalization 15.00%) | | | |
| 31.20% (scenario 29.04-33.30%) | | 51.41% (scenario 46.92-55.50%) | |
| **CKD - Base case VE** (rVE against infection 7.60%; rVE against hospitalization 8.40%) | | | |
| 29.11% (scenario 26.40–31.70%) | | 54.91% (scenario 51.01–58.49%) | |

**CKD:** chronic kidney disease; **CLD:** chronic lung disease; **CVD:** cardiovascular disease; **DM:** diabetes mellitus; **IC:** immune-compromised; **LB/UB:** lower bound/ upper bound

**Table S8. Vaccination coverage rates (%) (general population)**

| Age group | Sep23 | Oct23 | Nov23 | Dec23 | Jan24 | Feb24 | Mar24 | Apr24 | May24 | Jun24 | Jul24 | Aug24 |
|---|---|---|---|---|---|---|---|---|---|---|---|---|
| **Base case vaccination coverage** | | | | | | | | | | | | |
| 18-49 y | 0.64 | 3.71 | 8.20 | 9.50 | 12.24 | 12.50 | 12.50 | 12.50 | 12.50 | 12.50 | 12.50 | 12.50 |
| 50-64 y | 1.72 | 9.60 | 18.36 | 21.17 | 23.21 | 24.80 | 24.80 | 24.80 | 24.80 | 24.80 | 24.80 | 24.80 |
| ≥65 y | 3.08 | 17.10 | 31.64 | 33.88 | 38.94 | 39.80 | 39.80 | 39.80 | 39.80 | 39.80 | 39.80 | 39.80 |
| **Scenario: base case +10% coverage** | | | | | | | | | | | | |
| 18-49 y | 0.70 | 4.09 | 9.02 | 10.45 | 13.47 | 13.75 | 13.75 | 13.75 | 13.75 | 13.75 | 13.75 | 13.75 |
| 50-64 y | 1.89 | 10.56 | 20.19 | 23.28 | 25.54 | 27.28 | 27.28 | 27.28 | 27.28 | 27.28 | 27.28 | 27.28 |
| ≥65 y | 3.39 | 18.81 | 34.81 | 37.27 | 42.84 | 43.78 | 43.78 | 43.78 | 43.78 | 43.78 | 43.78 | 43.78 |
| **Scenario: Healthy people target rate of 70%** | | | | | | | | | | | | |
| 18-49 y | 3.58 | 20.80 | 45.92 | 53.20 | 68.56 | 70.00 | 70.00 | 70.00 | 70.00 | 70.00 | 70.00 | 70.00 |
| 50-64 y | 4.85 | 27.09 | 51.81 | 59.74 | 65.52 | 70.00 | 70.00 | 70.00 | 70.00 | 70.00 | 70.00 | 70.00 |
| ≥65 y | 5.42 | 30.08 | 55.65 | 59.59 | 68.49 | 70.00 | 70.00 | 70.00 | 70.00 | 70.00 | 70.00 | 70.00 |
| **Scenario: Two-dose vaccination** | | | | | | | | | | | | |
| 1st dose | | | | | | | | | | | | |
| 18-49 y | Same as base case | | | | | | | | | | | |

| | | | | | | | | | | | | |
|---|---|---|---|---|---|---|---|---|---|---|---|---|
| 50-64 y | | | | | | | | | | | | |
| ≥65 y | | | | | | | | | | | | |
| 2nd dose | | | | | | | | | | | | |
| 18-49 y | 0.00 | 0.00 | 0.00 | 0.00 | 0.00 | 0.00 | 0.00 | 0.00 | 0.00 | 0.00 | 0.00 | 0.00 |
| 50-64 y | 0.00 | 0.00 | 0.00 | 0.00 | 0.00 | 0.00 | 0.00 | 0.00 | 0.00 | 0.00 | 0.00 | 0.00 |
| ≥65 y | 0.00 | 0.00 | 0.00 | 0.00 | 0.00 | 0.00 | 7.10 | 10.00 | 10.00 | 10.00 | 10.00 | 10.00 |

**y:** year

**File S2. Indirect costs and QALYs**

Participation in the labor force was calculated from average employment rates in the US general population [13], adjusted to reflect decreased employment rates in high-risk populations. An RR of 0.55 was applied for CLD (based on COPD data [14]); an RR of 0.74 was applied for DM (based on Behavioral Risk Factor Surveillance System Survey (BRFSS) data for 2013 [15]); and a 28% decrease was applied for CVD [16]. The proportion of CKD patients in the labor force was calculated as a weighted average of employment rates [17] in the population with CKD by stage (distribution from USRDS [18]), assuming only stages 3 to 5 were working (i.e., excluding dialysis patients). Due to substantial heterogeneity in the IC population (e.g., employment rates in various IC populations ranged from 22.00% [19] to 77.30% [20]), the employment rate in IC patients was assumed to be 50% lower than in the general population, and this 50% decrease was applied to the age-stratified proportion of individuals in the US labor force[13].

The daily wage was estimated from the mean wage and working days reported for the US in 2023 [21, 22]. Lost productivity due to vaccine administration, outpatient care and hospitalization recovery was based on a previous US economic analysis for COVID-19 [12]. Lost productivity due to hospitalization was based on CDC data on the average length of stay of confirmed COVID-19 inpatient discharges [23]. For the IC population, lost productivity due to hospitalization was based on the average length of stay for the first COVID-19 hospitalization in IC patients [24]; and for CKD, lost productivity due to hospitalization [25] was adjusted to account for increased productivity losses reported in this group [26] (**Table S5**).

The number of working days lost due to severe long COVID was based on the most recent available data in the general population [27, 28]. For CLD, data on absenteeism for working patients with COPD [29] were adjusted by the RR for the number of days lost due to long COVID-19 in CLD patients [30]. For CVD, general population data [22] were adjusted by the decrease in income for CVD patients [16], and relative increase in the number of days lost due to long COVID-19 in patients with CVD [30]. For the IC population, number of working days lost due to severe long COVID was based on data for cancer with active treatment [31], and adjusted by the relative increase in the number of days lost due to long COVID in patients with cancer [30]. For

CKD, additional days lost calculated due to severe long COVID [17, 18, 30] assumed a comparable loss to patients with DM (**Table S5**).

QALYs/ utilities

The model applied baseline utility values [32] stratified by age group. Utility decrements associated with infections and hospitalization were from 5UM-CDC data [33]. Utility decrements associated with hospital re-admission, myocarditis, post-infection period, long COVID, and adverse events were based on a previous US economic analysis of COVID-19 in the general population [12], assumed to apply to high-risk populations due to lack of data (**Table S6**).

**File S3. Primary analysis results**

**Fig S1. A) Total healthcare and societal cost savings ($M), and B) patient-level cost savings ($) and ROI (payer/societal perspectives) with updated Moderna COVID-19 vaccination versus no updated vaccination**

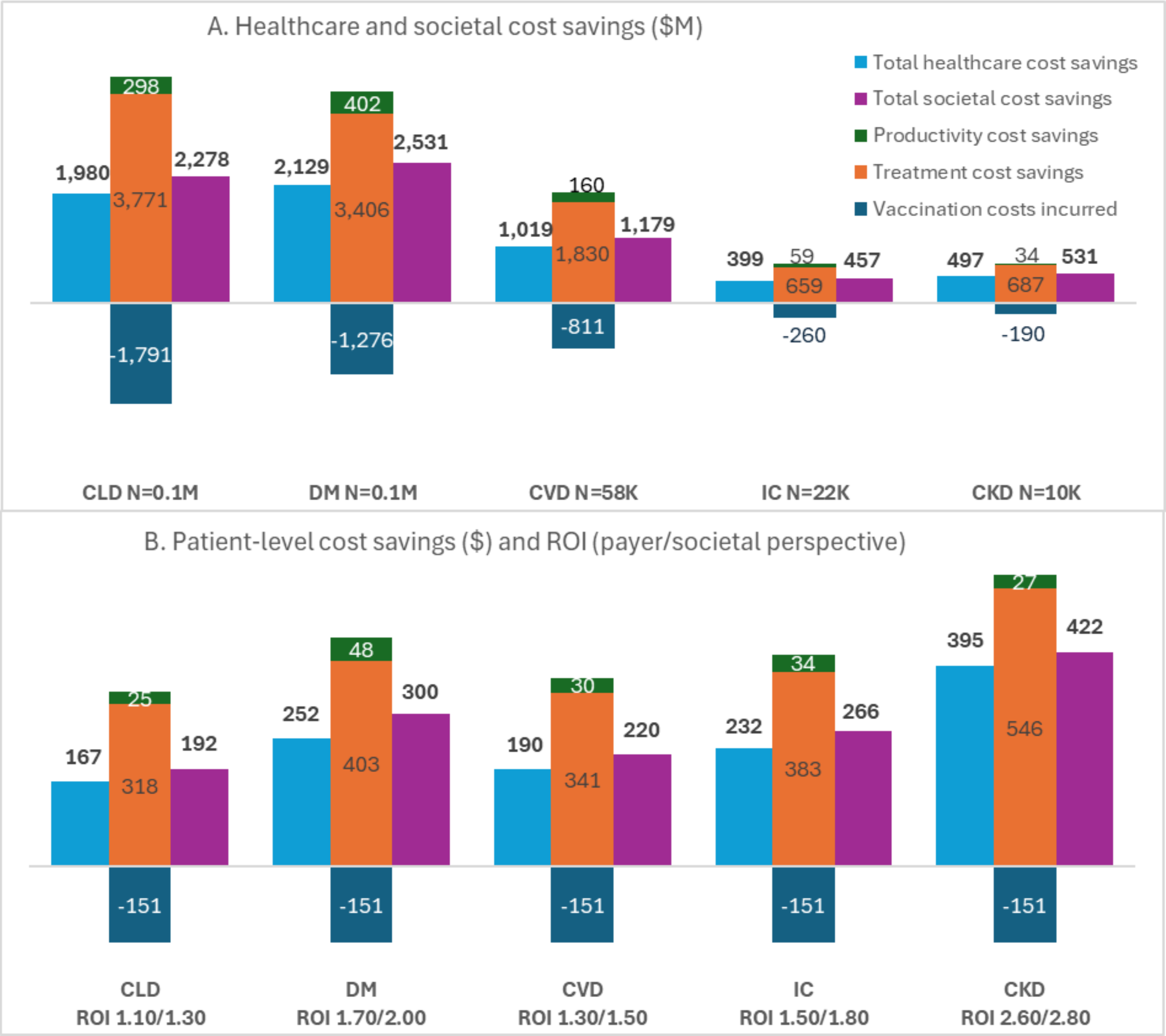


**CKD:** chronic kidney disease; **CLD:** chronic lung disease; **CVD:** cardiovascular disease; **DM:** diabetes mellitus; **IC:** immuno-compromised; **M:** million; **ROI:** return-on-investment

**Table S9. ICERs and ROIs with updated Moderna COVID-19 vaccination versus no updated vaccination (Healthcare payer and Societal perspectives)**

| | | **Updated Moderna COVID-19 vaccine** | **No updated vaccination** | **Difference** |
|---|---|---|---|---|
| **CLD** | Total QALYs lost | 1,267,930 | 1,428,672 | -160,742 |
| | Total costs (Healthcare), $ | 42,748,683,383 | 44,728,385,642 | -1,979,702,259 |
| | Total costs (Societal), $ | 46,989,284,107 | 49,266,941,919 | -2,277,657,812 |

| | | | | |
|---|---|---|---|---|
| | **ICER** | | | **Dominant** |
| | **ROI (patient-level, Healthcare)** | | | 1.10 |
| | **ROI (patient-level, Societal)** | | | 1.30 |
| **DM** | Total QALYs lost | 1,478,263 | 1,646,133 | -167,870 |
| | Total costs (Healthcare), $ | 40,065,273,678 | 42,194,428,399 | -2,129,154,721 |
| | Total costs (Societal), $ | 46,060,312,139 | 48,591,332,536 | -2,531,020,397 |
| | **ICER** | | | **Dominant** |
| | **ROI (patient-level, Healthcare)** | | | 1.70 |
| | **ROI (patient-level, Societal)** | | | 2.00 |
| **CVD** | Total QALYs lost | 672,410 | 760,600 | -88,191 |
| | Total costs (Healthcare), $ | 19,943,615,352 | 20,962,298,340 | -1,018,682,988 |
| | Total costs (Societal), $ | 22,299,388,634 | 23,478,117,070 | -1,178,728,436 |
| | **ICER** | | | **Dominant** |
| | **ROI (patient-level, Healthcare)** | | | 1.30 |
| | **ROI (patient-level, Societal)** | | | 1.50 |
| **IC** | Total QALYs lost | 337,257 | 370,216 | -32,959 |
| | Total costs (Healthcare), $ | 9,287,636,844 | 9,686,294,772 | -398,657,928 |
| | Total costs (Societal), $ | 10,338,726,629 | 10,796,130,732 | -457,404,103 |
| | **ICER** | | | **Dominant** |
| | **ROI (patient-level, Healthcare)** | | | 1.50 |
| | **ROI (patient-level, Societal)** | | | 1.80 |
| **CKD** | Total QALYs lost | 145,973 | 166,549 | -20,577 |
| | Total costs (Healthcare), $ | 6,025,268,254 | 6,521,907,134 | -496,638,880 |
| | Total costs (Societal), $ | 6,557,600,514 | 7,088,580,989 | -530,980,475 |
| | **ICER** | | | **Dominant** |
| | **ROI (patient-level, Healthcare)** | | | 2.60 |
| | **ROI (patient-level, Societal)** | | | 2.80 |

**Dominant ICER** signifies cost savings and QALYs saved with vaccination; **CKD:** chronic kidney disease; **CLD:** chronic lung disease; **CVD:** cardiovascular disease; **DM:** diabetes mellitus; **IC:** immuno-compromised; **ICER:** incremental cost-effectiveness ratio; **QALY:** quality-adjusted life-year; **ROI:** return on investment

### Fig S2. Scenario analysis results: Tornado diagram for healthcare cost savings and QALYs saved – DM population

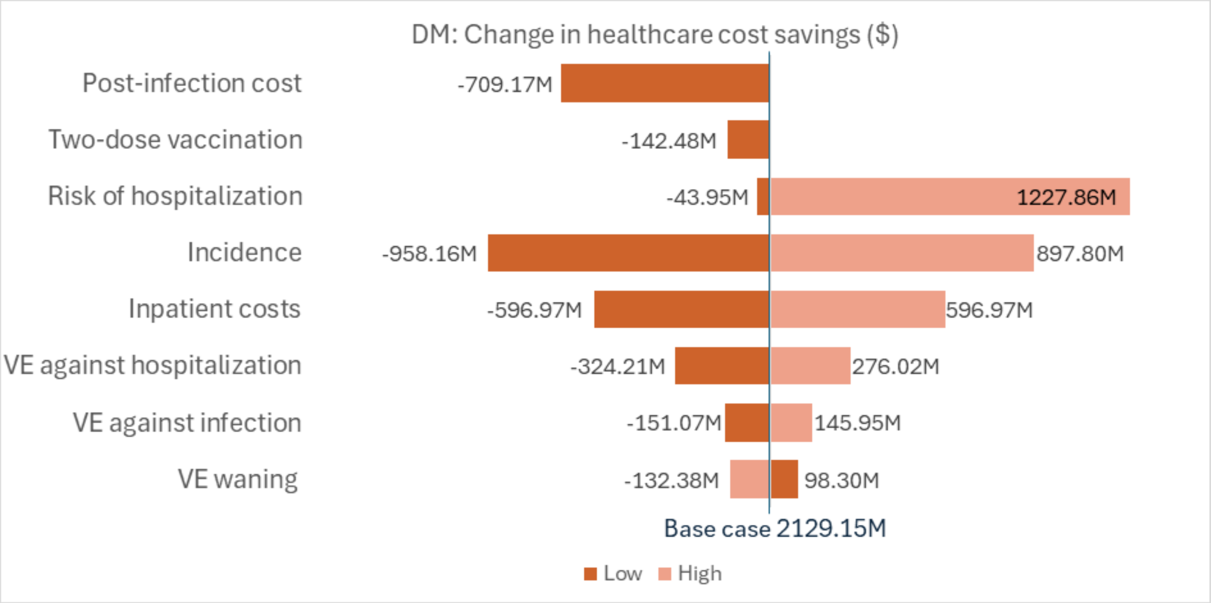
DM: Change in healthcare cost savings ($)
Post-infection cost
Two-dose vaccination
Risk of hospitalization
Incidence
Inpatient costs
VE against hospitalization
VE against infection
VE waning
-709.17M
-142.48M
-43.95M
1227.86M
-958.16M
897.80M
-596.97M
596.97M
-324.21M
276.02M
-151.07M
145.95M
-132.38M
98.30M
Base case 2129.15M
Low
High

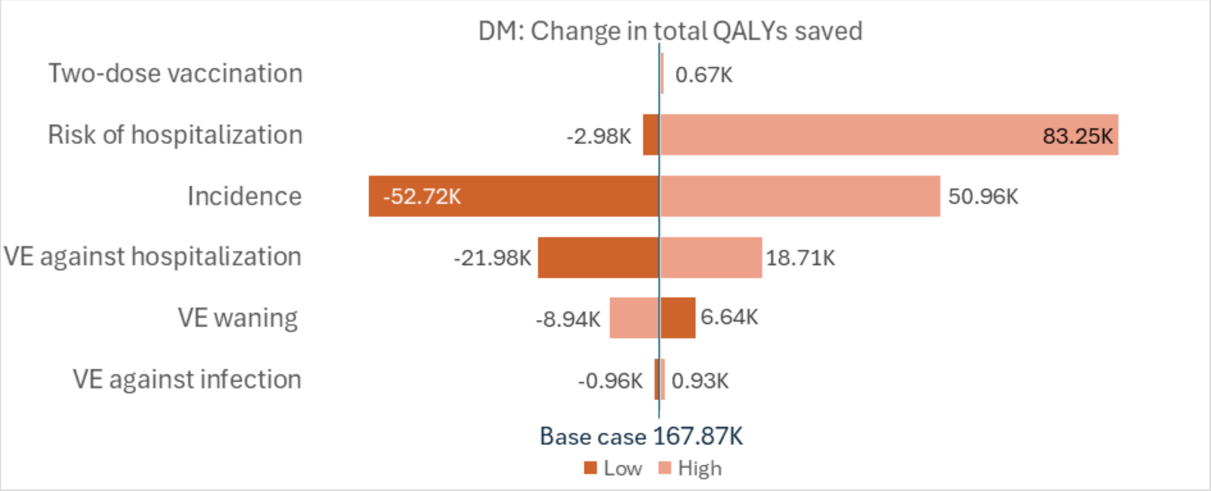
DM: Change in total QALYs saved
Two-dose vaccination
Risk of hospitalization
Incidence
VE against hospitalization
VE waning
VE against infection
0.67K
-2.98K
83.25K
-52.72K
50.96K
-21.98K
18.71K
-8.94K
6.64K
-0.96K
0.93K
Base case 167.87K
Low
High

**Fig S3. Scenario analysis results: Tornado diagram for healthcare cost savings and QALYs saved – CVD population**

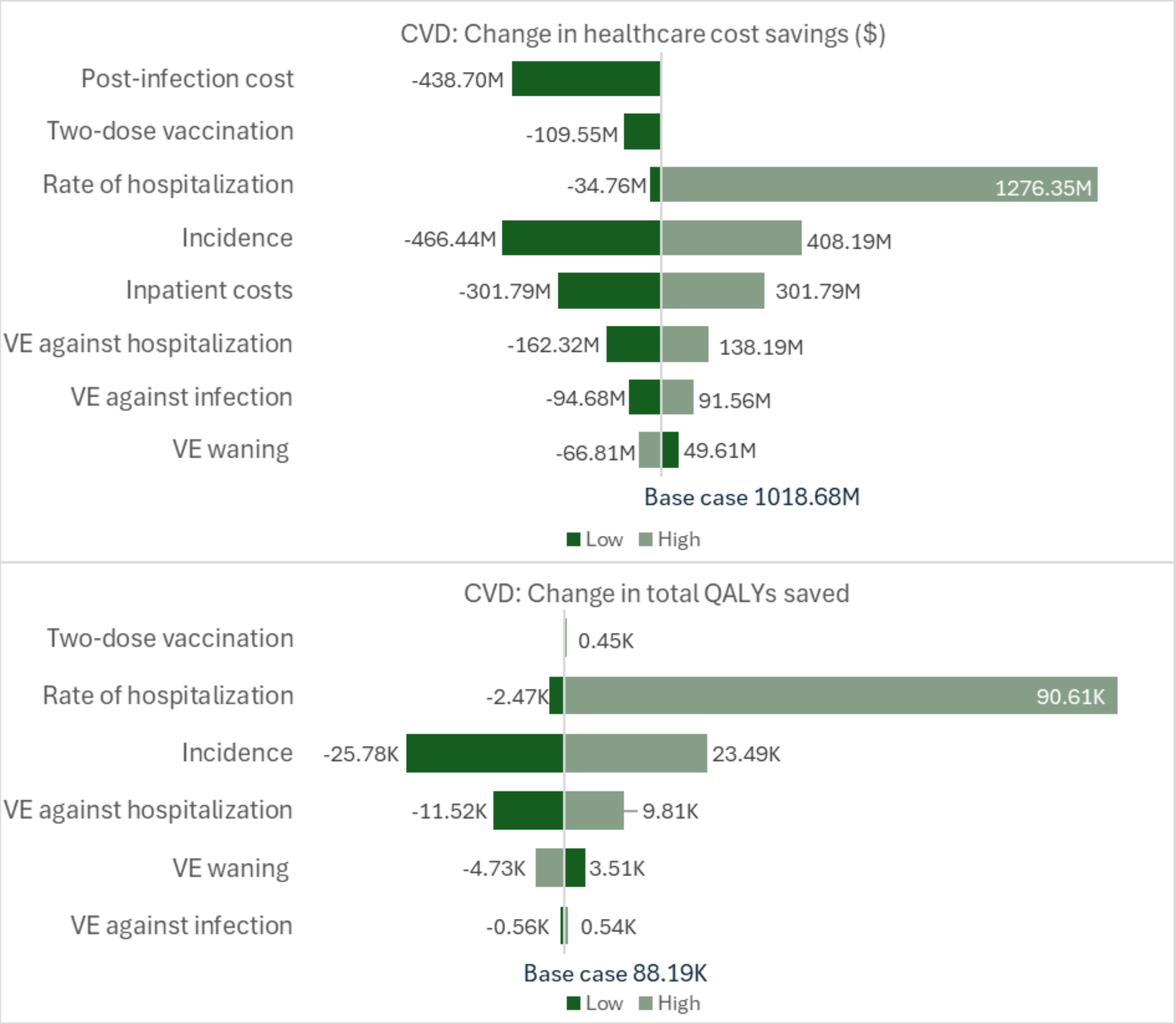

**Fig S4. Scenario analysis results: Tornado diagram for healthcare cost savings and QALYs saved – IC population**

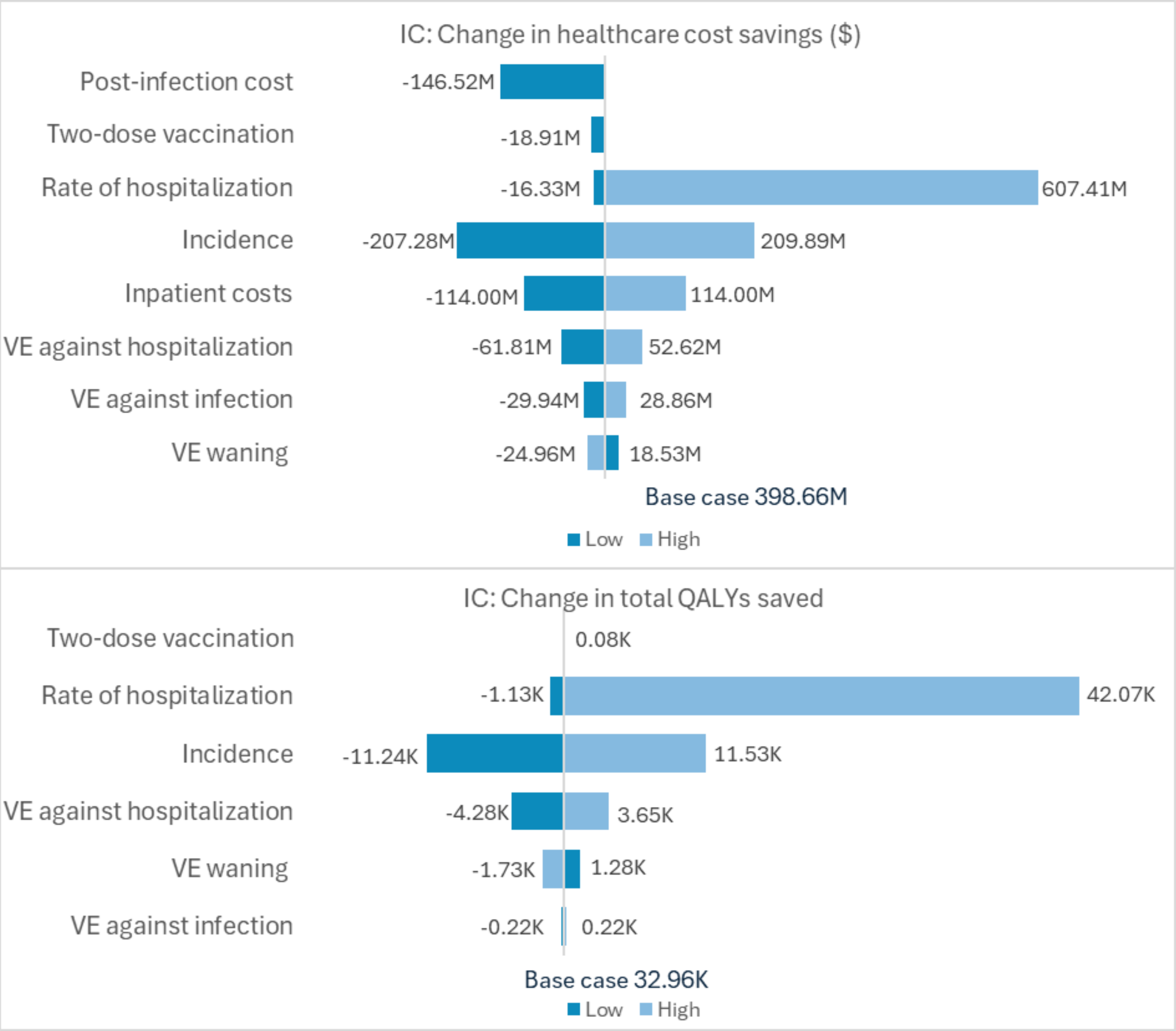

**Fig S5. Scenario analysis results: Tornado diagram for healthcare cost savings and QALYs saved – CKD population**

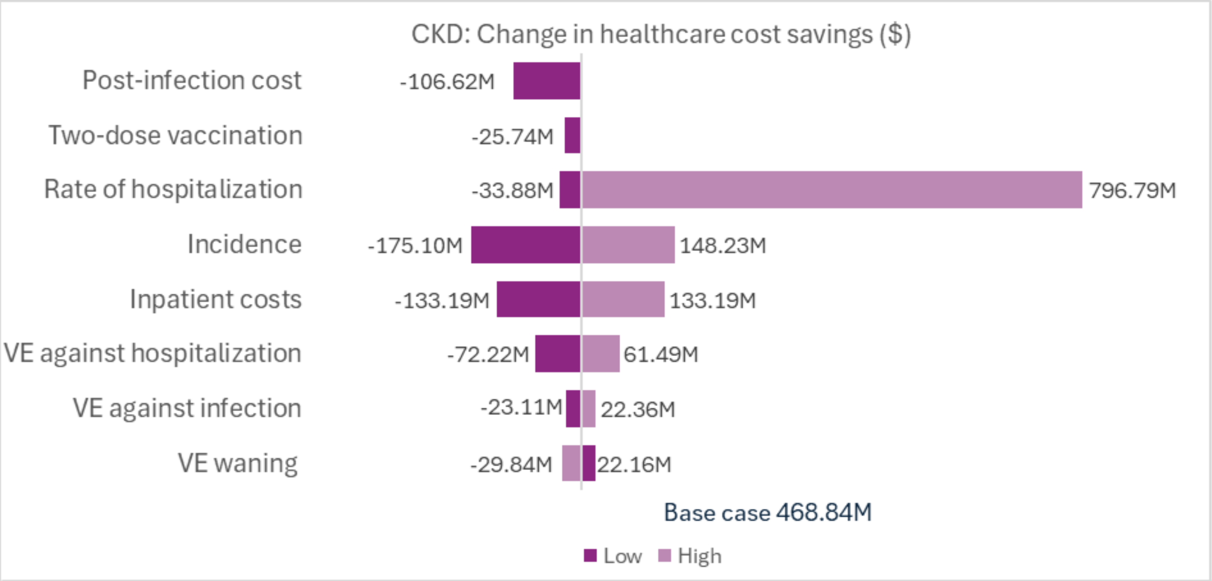


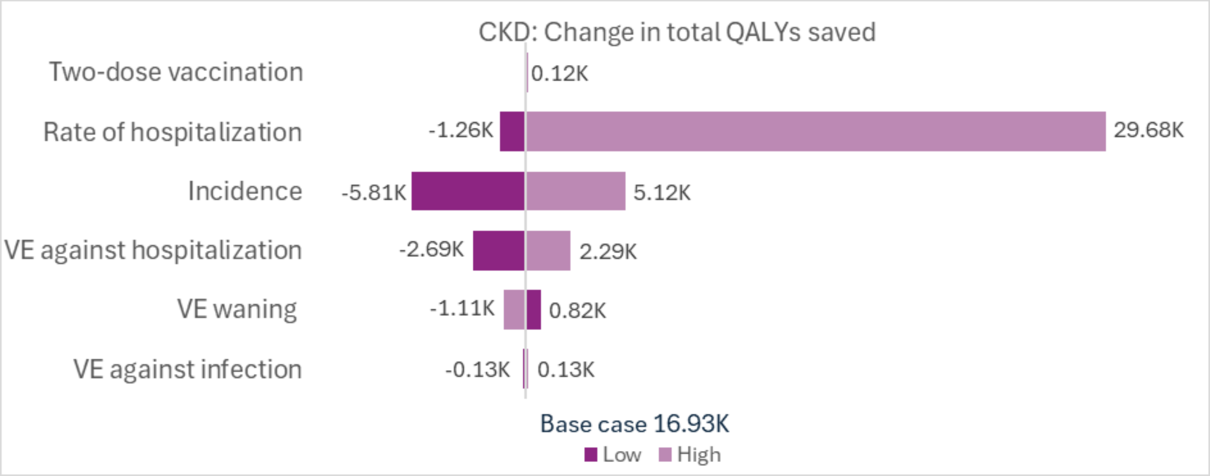

**File S4. Secondary analysis results**

Results for Moderna's versus Pfizer/BioNTech's updated vaccination

**Table S10. ICERs with Moderna's versus Pfizer/BioNTech's updated COVID-19 vaccination (Healthcare payer and Societal perspectives)**

| | | Updated Moderna COVID-19 vaccine | Updated Pfizer/BioNTech COVID-19 vaccine | Difference |
|---|---|---|---|---|
| **CLD** | Total QALYs lost | 1,267,930 | 1,284,419 | -16,489 |
| | Total costs (Healthcare), $ | 42,748,683,383 | 43,118,842,484 | -370,159,101 |
| | Total costs (Societal), $ | 46,989,284,107 | 47,393,625,182 | -404,341,074 |
| | **ICER** | | | **Dominant** |
| **DM** | Total QALYs lost | 1,478,263 | 1,500,829 | -22,567 |
| | Total costs (Healthcare), $ | 40,065,273,678 | 40,498,847,181 | -433,573,503 |
| | Total costs (Societal), $ | 46,060,312,139 | 46,552,299,237 | -491,987,099 |
| | **ICER** | | | **Dominant** |
| **CVD** | Total QALYs lost | 672,410 | 684,372 | -11,963 |
| | Total costs (Healthcare), $ | 19,943,615,352 | 20,246,610,149 | -302,994,797 |
| | Total costs (Societal), $ | 22,299,388,634 | 22,630,627,166 | -331,238,533 |
| | **ICER** | | | **Dominant** |
| **IC** | Total QALYs lost | 337,257 | 341,697 | -4,440 |
| | Total costs (Healthcare), $ | 9,287,636,844 | 9,377,607,912 | -89,971,068 |
| | Total costs (Societal), $ | 10,338,726,629 | 10,437,794,549 | -99,067,920 |
| | **ICER** | | | **Dominant** |
| **CKD** | Total QALYs lost | 145,973 | 147,560 | -1,588 |
| | Total costs (Healthcare), $ | 6,025,268,254 | 6,099,232,025 | -73,963,771 |
| | Total costs (Societal), $ | 6,557,600,514 | 6,635,638,923 | -78,038,409 |
| | **ICER** | | | **Dominant** |

**Dominant ICER** signifies cost savings and QALYs saved with vaccination; **CKD:** chronic kidney disease; **CLD:** chronic lung disease; **CVD:** cardiovascular disease; **DM:** diabetes mellitus; **IC:** immuno-compromised; **ICER:** incremental cost-effectiveness ratio; **QALY:** quality-adjusted life-year

For the comparison against Pfizer/BioNTech's updated vaccine, scenarios assessed the impact of varying the rVE estimates, and of increasing coverage.

**Table S11. Scenario analyses: Healthcare cost savings, QALYs saved, and ICERs with Moderna's versus Pfizer/BioNTech's updated COVID-19 vaccination (Healthcare payer perspective)**

| **CLD** | **Moderna** | **Pfizer/BioNTech** | **Difference** |
|---|---|---|---|
| **Scenario +10% increase in VCR** | | | |
| Total QALYs lost | 1,251,856 | 1,269,994 | -18,138 |
| Total costs (Healthcare), $ | 42,550,713,157 | 42,957,888,168 | -407,175,012 |
| ICER, $ per QALY / Conclusion | | | **Dominant** |

| | | | |
|---|---|---|---|
| **Scenario VCR 70%** | | | |
| Total QALYs lost | 1,030,360 | 1,071,153 | -40,793 |
| Total costs (Healthcare), $ | 40,031,764,231 | 40,877,477,422 | -845,713,191 |
| ICER, $ per QALY / Conclusion | | | **Dominant** |
| **Scenario Lower bound of 95%CI for rVE estimate** | | | |
| Total QALYs lost | 1,267,930 | 1,274,267 | -6,338 |
| Total costs (Healthcare), $ | 42,748,683,383 | 42,855,215,986 | -106,532,604 |
| ICER, $ per QALY / Conclusion | | | **Dominant** |
| **Scenario Higher bound of 95%CI for rVE estimate** | | | |
| Total QALYs lost | 1,267,930 | 1,295,270 | -27,340 |
| Total costs (Healthcare), $ | 42,748,683,383 | 43,393,054,237 | -644,370,854 |
| ICER, $ per QALY / Conclusion | | | **Dominant** |
| **DM** | **Moderna** | **Pfizer/BioNTech** | **Difference** |
| **Scenario +10% increase in VCR** | | | |
| Total QALYs lost | 1,461,476 | 1,486,299 | -24,824 |
| Total costs (Healthcare), $ | 39,852,358,206 | 40,329,289,059 | -476,930,853 |
| ICER, $ per QALY / Conclusion | | | **Dominant** |
| **Scenario VCR 70%** | | | |
| Total QALYs lost | 1,181,176 | 1,243,580 | -62,404 |
| Total costs (Healthcare), $ | 36,487,255,738 | 37,610,926,891 | -1,123,671,153 |
| ICER, $ per QALY / Conclusion | | | **Dominant** |
| **Scenario Lower bound of 95%CI for rVE estimate** | | | |
| Total QALYs lost | 1,478,263 | 1,490,202 | -11,940 |
| Total costs (Healthcare), $ | 40,065,273,678 | 40,271,096,269 | -205,822,591 |
| ICER, $ per QALY / Conclusion | | | **Dominant** |
| **Scenario Higher bound of 95%CI for rVE estimate** | | | |
| Total QALYs lost | 1,478,263 | 1,512,131 | -33,868 |
| Total costs (Healthcare), $ | 40,065,273,678 | 40,739,798,307 | -674,524,629 |
| ICER, $ per QALY / Conclusion | | | **Dominant** |
| **CVD** | **Moderna** | **Pfizer/BioNTech** | **Difference** |
| **Scenario +10% increase in VCR** | | | |
| Total QALYs lost | 663,591 | 676,749 | -13,159 |
| Total costs (Healthcare), $ | 19,841,747,054 | 20,175,041,330 | -333,294,277 |
| ICER, $ per QALY / Conclusion | | | **Dominant** |
| **Scenario VCR 70%** | | | |
| Total QALYs lost | 544,717 | 573,921 | -29,204 |
| Total costs (Healthcare), $ | 18,530,923,511 | 19,212,137,622 | -681,214,110 |
| ICER, $ per QALY / Conclusion | | | **Dominant** |
| **Scenario Lower bound of 95%CI for rVE estimate** | | | |
| Total QALYs lost | 672,410 | 679,329 | -6,920 |
| Total costs (Healthcare), $ | 19,943,615,352 | 20,127,092,216 | -183,476,864 |
| ICER, $ per QALY / Conclusion | | | **Dominant** |
| **Scenario Higher bound of 95%CI for rVE estimate** | | | |
| Total QALYs lost | 672,410 | 689,764 | -17,355 |
| Total costs (Healthcare), $ | 19,943,615,352 | 20,369,442,608 | -425,827,256 |
| ICER, $ per QALY / Conclusion | | | **Dominant** |
| **IC** | **Moderna** | **Pfizer/BioNTech** | **Difference** |
| **Scenario +10% increase in VCR** | | | |
| Total QALYs lost | 333,961 | 338,845 | -4,884 |
| Total costs (Healthcare), $ | 9,247,771,052 | 9,346,739,226 | -98,968,174 |
| ICER, $ per QALY / Conclusion | | | **Dominant** |
| **Scenario VCR 70%** | | | |
| Total QALYs lost | 267,831 | 281,603 | -13,772 |
| Total costs (Healthcare), $ | 8,457,680,341 | 8,726,082,269 | -268,401,929 |

| | | | |
|---|---|---|---|
| ICER, $ per QALY / Conclusion | | | **Dominant** |
| **Scenario Lower bound of 95%CI for rVE estimate** | | | |
| Total QALYs lost | 337,257 | 339,190 | -1,934 |
| Total costs (Healthcare), $ | 9,287,636,844 | 9,324,570,633 | -36,933,789 |
| ICER, $ per QALY / Conclusion | | | **Dominant** |
| **Scenario Higher bound of 95%CI for rVE estimate** | | | |
| Total QALYs lost | 337,257 | 344,437 | -7,181 |
| Total costs (Healthcare), $ | 9,287,636,844 | 9,434,407,244 | -146,770,400 |
| ICER, $ per QALY / Conclusion | | | **Dominant** |
| **CKD** | **Moderna** | **Pfizer/BioNTech** | **Difference** |
| **Scenario +10% increase in VCR** | | | |
| Total QALYs lost | 143,915 | 145,661 | -1,746 |
| Total costs (Healthcare), $ | 5,975,604,366 | 6,056,964,514 | -81,360,148 |
| ICER, $ per QALY / Conclusion | | | **Dominant** |
| **Scenario VCR 70%** | | | |
| Total QALYs lost | 119,028 | 122,675 | -3,647 |
| Total costs (Healthcare), $ | 5,425,902,930 | 5,582,147,902 | -156,244,973 |
| ICER, $ per QALY / Conclusion | | | **Dominant** |
| **Scenario Lower bound of 95%CI for rVE estimate** | | | |
| Total QALYs lost | 145,973 | 146,159 | -186 |
| Total costs (Healthcare), $ | 6,025,268,254 | 6,046,327,448 | -21,059,194 |
| ICER, $ per QALY / Conclusion | | | **Dominant** |
| **Scenario Higher bound of 95%CI for rVE estimate** | | | |
| Total QALYs lost | 145,973 | 149,077 | -3,104 |
| Total costs (Healthcare), $ | 6,025,268,254 | 6,155,051,184 | -129,782,930 |
| ICER, $ per QALY / Conclusion | | | **Dominant** |

**Dominant ICER** signifies cost savings and QALYs saved; **CI:** confidence interval; **CKD:** chronic kidney disease; **CLD:** chronic lung disease; **CVD:** cardiovascular disease; **DM:** diabetes mellitus; **IC:** immuno-compromised; **ICER:** incremental cost-effectiveness ratio; **QALY:** quality-adjusted life-year; **rVE:** relative vaccine efficacy

33. 5UM-CDC data source: Coronavirus Household Evaluation and Respiratory Testing (CHEaRT) and Prospective Assessment of COIVD 19 in a Community (PACC), unpublished data.
